# Early Precursors of Literacy Development in Simultaneous Bilinguals: A Systematic Review and Meta-Analysis

**DOI:** 10.1101/2021.08.18.21262243

**Authors:** Insiya Bhalloo, Monika Molnar

## Abstract

**Purpose:** Literacy precursors are cognitive, linguistic, and oral-language skills that predict future reading skills in children as young as 4 years. Speech-language pathologists and educators utilize these precursors as assessment tools to identify children at risk for reading difficulties. Most current tools are developed based on monolinguals (predominantly in English), despite the significant percentage of bilinguals globally. As such, bilingual children are typically assessed on tools developed for monolinguals in research and clinical settings. Despite this common practice, there is a lack of comprehensive synthesis on whether these precursors are a reliable indicator of reading skills in bilingual children. Our paper examines whether literacy precursors commonly used with monolinguals are associated with literacy development in simultaneous bilinguals.

**Method:** Following PRISMA and Cochrane guidelines, our review includes four databases (LLBA, ERIC, MLA, and PsycINFO), in addition to gray-literature and manual reference-list searches. To control for age of acquisition and language dominance variability, we included typically-developing simultaneous bilinguals exposed to both languages before age 3 (N=5,942). We analyzed reported statistical associations between code-related or oral-language precursors and reading outcome measures, using correlational meta-analyses.

**Results:** The 41 reports, that met inclusion/exclusion criteria, were published between 1977-2022. The average age at assessment was 7;5 years (range: 3;0 - 11;0 years), with children speaking over 21 bilingual language combinations. Our meta-analysis demonstrated significant within-language correlations and cross-language transfer effects for code-related (e.g., phonological awareness) and oral-language (e.g., vocabulary, morphological awareness) precursors. Semantic awareness, however, was not a reliable predictor in bilinguals.

**Conclusions:** Phonological awareness and vocabulary measures – even if originally developed for monolingual children - can form a meaningful component of early literacy assessment in simultaneous bilingual children: these precursors may be used as assessment tools across heritage and societal languages in research and clinical practice. Future research suggestions within this domain are also discussed.

## Introduction

Globally, 56% of children do not meet age-appropriate reading levels (United Nations Educational, Scientific and Cultural Organization [UNESCO], 2017). Age-appropriate literacy skills (i.e., reading and writing) are linked with long-term academic and socio-economic success in both monolingual and bilingual children (Ritchie & Bates, 2013). Bilingual children — who make up a substantial number of new students entering the school system every year (Ryan, 2013) — however, are disproportionately under/over-identified for reading difficulties at preschool and Grade 1 levels, compared to their monolingual peers (Altman et al., 2012; Samson & Lesaux, 2009).

Could this be because current literacy assessment tools have been predominantly developed for monolinguals? The current synthesis and meta-analysis are an initial step towards addressing this complex question. There are a multitude of other factors, beside the lack of linguistically- and culturally-appropriate assessment tools, potentially contributing to the misidentification of bilingual children with language and reading difficulties. These include limited bilingual speech-language pathologists, lack of bilingual performance norms, unidentified early literacy milestones and indicators of reading difficulties, and inconsistent assessment practices for languages other than English (American-Speech-Language-Hearing Association [ASHA], 2021; Goodrich et al., 2021; Teoh et al., 2018). It is beyond the current review to investigate all of these factors. Here, we focus on the effectiveness of assessing *early/emergent literacy skills* to predict future reading abilities in bilinguals. These bilingual assessments are typically conducted with tools established with monolingual populations.

To identify children at risk for future reading difficulties, speech-language pathologists (SLPs) and educators often assess *emergent literacy skills* (Puolakanaho et al., 2007). Emergent literacy refers to aspects of literacy that develop prior to receiving formal reading instruction in school settings; this includes *literacy precursors*. Literacy precursors are a subset of linguistic and cognitive skills, developing during preschool age, that facilitate word-level decoding, word recognition, and future text reading comprehension (Scarborough, 2001). Previous systematic reviews and meta-analyses in monolingual English-speaking children have categorized literacy precursors into *code-related, oral-language*, and *domain-general cognitive* skills (García & Cain, 2014; Hjetland et al., 2017, 2020; National Early Literacy Panel [NELP], 2008). Literacy precursors are utilized as an indicator of typical reading development in early reading programs and specifically at preschool - Grade 2 levels, and for continued monitoring throughout the child’s reading development trajectory (Georgiou, 2017; Savage et al., 2018).

Code-related, oral-language, and domain-general cognitive skills are also featured in prominent reading models. *Code-related skills* include phonological awareness, rapid automatized naming (RAN), and letter knowledge. Several models highlight the importance of code-related skills for word decoding (WD) and reading comprehension (RC) at the early stages of learning to read (Simple View of Reading or SVR; Hoover & Gough, 1990; Scarborough’s (2001) Rope Model; McKenna & Stahl’s (2009) Modified Cognitive Model of Reading Comprehension). These skills enable children to understand systematic relationships between oral and written language and acquire symbol-sound (i.e., letter-phoneme) mapping principles crucial for transitioning from accurate WD to later fluent word recognition (Hjetland et al., 2017, 2020; Lonigan et al., 2018). Children at higher grades, with already established word recognition skills, also greatly rely on cognitive and oral-language skills to recognize words and comprehend complex text (Fletcher-Campbell et al., 2009). *Oral-language skills* involve understanding and producing spoken language, and include vocabulary and grammatical awareness. Dual route and parallel-distributed-processing (PDP) connectionist reading models, that primarily describe proficient reading in older children and adults, emphasize both code-related and oral-language skills for word reading strategies (Coltheart & Rastle, 1994; Seidenberg & McClelland, 1989).

In addition, reading and comprehension require *domain-general cognitive skills*. These skills are demonstrated by non-linguistic memory, reasoning, and ‘intelligence’ tasks, and allow children to comprehend text and conduct active monitoring, simultaneous recall, manipulation and integration of incoming and background information (Tighe et al., 2015). Domain-general cognitive precursors are also important for monolingual WD and fluent word recognition (Bhattacharya & Ehri, 2004; Treiman et al., 1990). Further, bilingual word recognition involves inhibition of non-target linguistic information from related languages, which requires domain-general cognitive processes (Bilingual Interactive Activation Plus model; BIA+; Diependaele et al., 2010).

In terms of bilinguals, most reading models address how proficient adult readers are affected by being able to read in two languages. Apart from the Linguistic Interdependence Hypothesis (LIH; Cummins, 1979) and its extension - the Interactive Transfer Framework (Chung et al., 2019), Script Dependent Hypothesis (SDH; Geva & Siegel, 2000), and Psycholinguistic Grain Size Theory (PGST; Ziegler & Goswami, 2005; 2006), these models are based on European alphabetic languages, and primarily English monolinguals (Daniels & Share, 2018; Nation, 2019; Share, 2008). The LIH, Interactive Transfer Framework, SDH, and PGST highlight influences of heritage-language vocabulary as well as orthographic and phonological features on reading development milestones, including cross-language literacy skill transfer (Bialystok et al., 2005). Specifically, the PGST compares the relationship between phonological awareness measures (a type of code-related literacy precursor), and word-level reading performance across orthographic depth (Ziegler & Goswami, 2005; 2006). For instance, Greek, Finnish, German, Italian, and Spanish readers of orthographically-transparent scripts demonstrate higher decoding scores and fluent reading abilities at an earlier age, compared to English readers (Aro & Wimmer, 2003; Seymour et al., 2003). This is due to consistent grapheme-phoneme correspondences (GPCs) at the phoneme grain size^1^ in orthographically-transparent scripts, which facilitate acquisition of symbol-sound mapping principles and use of phonological decoding-based initial reading strategies in line with dual-route models. In comparison, children learning to read in opaque orthographies, such as English, rely on multiple sub-lexical phonological and lexical reading strategies to recognize larger onset-rime grain sizes and decipher inconsistent mappings at the phoneme grain size level (Seymour et al., 2003; Ziegler & Goswami, 2006).

As highlighted above, reading can be influenced by different factors across bilinguals and monolinguals; yet research on bilingual reading is scarce. This lack of linguistic diversity in child literacy research influences clinical practice and vice versa: linguistically-appropriate assessments and literacy precursor performance norms are primarily available to support monolingual children (mostly in English). Here, we take a look at the currently existing research on bilingual children and evaluate whether literacy precursors – as measured by assessment tools – perform similarly in monolingual and bilingual children.

## Current Study

Assessment professionals with a bilingual caseload and researchers investigating bilingual development often have no choice, but to use assessment tools developed for monolinguals. This is due to the lack of resources specific to bilinguals (Gillam et al., 2013; Rose at al., 2022). In this systematic review and meta-analyses, we examine whether code-related, oral-language, and domain-general cognitive literacy precursors commonly used with monolinguals also correlate with early reading skills in bilinguals. We evaluate whether these precursors predict reading and comprehension at the word/non-word and text levels. This is because in our population of interest, which is primary school-aged children (i.e., between 4-12 years; preschool - Grade 7), word decoding is the most dominant skill (Scarborough, 2001). This skill is commonly used as an indicator of reading ability at this age (e.g., at 4-6 years), while emerging readers are still developing their word-level reading accuracy and fluency skills (Seymour et al., 2003). Strong decoders become future proficient readers, and then rely on oral-language and higher-level cognitive skills to rapidly recognize (e.g., at 6-8 years) and comprehend (e.g., at 8-12 years) text at later grade levels (Hernandez, 2011).

Also relevant to the current review is that bilingualism is not a categorical but a continuous variable. Bilingual children represent a heterogenous population in terms of the age of acquisition (AoA) and proficiency levels of their languages, in addition to the frequency/context of their exposure to the given languages. To control for this large variability in the bilingual experience, here, we investigate precursors of literacy development in *simultaneous bilingual children*. Simultaneous bilinguals acquire both languages prior to 3 years, and before formal literacy instruction (Patterson, 2002). AoA is a common categorization factor within bilinguals, as it has been shown to affect bilinguals’ oral-language and domain-general cognitive skills (e.g., Blom et al., 2014; Kuo et al., 2016). For instance, simultaneous bilinguals may demonstrate higher oral-language and passage RC scores, as compared to age-matched sequential bilinguals who learnt their second language after age 3 (Davison & Hammer, 2012; Gross et al., 2014). It should be noted there is valuable literature on sequential bilinguals and ‘second-language learners’ in terms of literacy precursors (e.g., August & Shanahan, 2006; Lesaux & Geva, 2006) – however, this segment of the bilingual population is beyond the scope of the current review. Here, we focus on simultaneous bilinguals – a considerable segment of the bilingual population, to ensure that comparisons in our review are meaningful and confound effects (e.g., due to differences in AoA, proficiency levels, etc.) are minimized.

## Objectives

This systematic review and meta-analyses examine whether: code-related (i.e., phonological awareness, letter knowledge, RAN), oral-language (i.e., vocabulary, syntactic awareness, morphological awareness), and domain-general cognitive (i.e., non-linguistic memory and intelligence) literacy precursors, or other precursors have been assessed and found to be associated with reading skills in one or both spoken languages of typically-developing, hearing, simultaneous bilingual children. In terms of the reading measures, we focus on word/non-word, sentence, and passage/text-level reading accuracy, fluency, and comprehension. As this paper focuses on pre-literacy skills in typically-developing bilingual children, our review and meta-analyses covers *assessment* studies only; intervention studies are beyond the scope.

## Method

### Search and Information Sources

We searched four databases, indexing language and literacy literature; these are Linguistics and Language Behavior Abstracts (LLBA), Educational Resources Information Center (ERIC), MLA International Bibliography, and PsycINFO (ProQuest). We conducted the latest search on February 22^nd^, 2023. The target literacy precursors were searched with the two outcome measures to find studies discussing at least one precursor in relation to word and/or text-level reading or comprehension. Search terms and syntax were determined in consultation with a librarian. Searches included peer-reviewed and gray literature, and unpublished research callouts. Further, we conducted a Google Scholar and manual reference search of studies. The Supplementary Tables (S1, S2, and S3) includes the detailed database search strategy, search syntax, and terms.

### Study Selection

Two independent reviewers used *Covidence* and *Excel* to screen each study based on inclusion, exclusion, and critical appraisal eligibility criteria at title & abstract and full-text levels. An inter-rater reliability (IRR) of 79.5% was achieved at full-text screening, in accordance with dichotomous ‘Yes/No’ IRR recommendations between 75%-90% for reviews (Graham et al., 2012). To ensure inclusion of appropriate studies, we added a ‘Maybe’ option which lowered our IRR. A third reviewer was consulted to reconcile reviewer differences and discuss studies rated as ‘Maybe’. We achieved 91.2% IRR after discussing and re-rating these studies. Most (i.e., ∼90%) differences were related to verifying simultaneous bilingual status, due to limited reported language background information and studies merging bilingual and monolingual groups in statistical analyses.

### Eligibility Criteria

Our inclusion criteria were as follows:

1. Assessed precursor skills in relation to reading-based literacy outcome measures, including, word/non-word and text-level reading accuracy, fluency, and/or comprehension;
2. Assessed typically-developing and hearing simultaneous bilinguals ≤ 12 years, and/or multilinguals and/or included age-matched typically-developing bilingual controls, who do not require and have not previously received language or literacy intervention;
3. Assessed simultaneous bilinguals, who were born or immigrated prior to 3 years to a country where the heritage language was not an official language and had sufficient exposure to the heritage language at home and school/societal language in wider community, even if the societal language was not the home language; or were exposed to both as home languages. Studies with monolingual or sequential bilingual groups were included if these also separately assessed and analyzed simultaneous bilingual groups.^2^

We excluded studies that met at least one of the following:

1. Non-primary (e.g., reviews) and case studies;
2. Non-English studies with inaccessible translations;
3. Studies on bimodal bilingual populations (i.e., with signed-spoken language combinations);
4. Intervention studies, that did not also assess precursor skills in typically-developing children prior to any intervention;
5. Studies only analyzing precursors in relation to other precursors, not reading outcomes;
6. Studies only assessing outcomes other than word, sentence or passage/text-level reading;
7. Analyses not separating non-typically developing, deaf or sequential bilinguals/dual language learners and monolinguals from typically-developing and hearing simultaneous bilinguals. ***Critical Appraisal***. To account for risk of bias, included studies met all criteria from an adapted Critical Appraisal Checklist for Quasi-Experimental Studies (Joanna Briggs Institute, 2017):

1. ***Clear Cause and Effect:*** Examined whether precursors were associated with outcomes;
2. ***Reliable Assessment and Outcome Measures:*** Described literacy assessment measures;
3. ***Appropriate Statistical Analyses:*** Used appropriate statistical tests;
4. ***Attrition Follow-Up Procedures in Longitudinal Studies:*** Accounted for participant attrition.
5. ***Additional Criteria – Sufficient Language and Socio-Economic Status (SES) Background:*** Specified adequate language information such as age of acquisition (AoA) to determine simultaneous bilingual status. We only included studies that reported or accounted for SES (see Data Items below), as bilingual experience and SES influence literacy skills (Meir & Armon-Lotem, 2017).

### Data Items

The following data items were extracted from the included studies indicated in Supplemental Table S4: *study citation, participant demographics* (i.e., sample size and participant groupings, age at evaluation, gender, place of birth or age at immigration, and language status in assessment country), *language background* (e.g., spoken language combinations, AoA, and degree/duration of language exposure and usage), *parental SES* (e.g., income, education, and home/school neighbourhood), *assessed literacy precursor and outcome measures* (i.e., type, measure and language(s) of assessment, and whether assessed in one or both languages), and *statistical analyses* (i.e., reported significant or non-significant associations between precursors and outcomes assessed in one or both languages, and correlation coefficients).

### Analyses

In our summary synthesis, *precursor-outcome associations* are reported statistically significant or non-significant relationships between a given literacy precursor and reading outcome measure. We categorize coefficients as weak (*r* < 0.4), moderate (*r*=0.4-0.6), or strong (*r*=0.7-0.9), as well as specify statistical significance as *p ≤* 0.05 for our random-effects correlational meta-analyses and as *p ≤* 0.1 for subgroup analyses in line with Cochrane systematic reviews (Dancey & Reidy, 2007; Richardson et al., 2019). The meta-analyses were conducted on correlation coefficients, as only 6/41 studies reported standardized mean difference (SMD or Cohen’s d) measures of effect size (see S4 for statistical methods). Following Cochrane Collaboration (2011)’s recommendations to account for statistical independence violations, we selected a single reported correlation coefficient for studies reporting multiple correlation coefficients across literacy precursor type.

We conducted random-effects correlational meta-analyses for precursors reporting a minimum of five independent correlation coefficients (Jackson & Turner, 2017), to account for between-study variation in sample size and measures. These include *code-related* (phonological awareness) and *oral-language/grammar* skills (vocabulary, morphological awareness) in relation to word/non-word reading, and for vocabulary and decoding in relation to text comprehension. To account for potential language effects on reading development (e.g., type of orthographic system; degree of orthographic depth), we conducted separate meta-analyses for precursors across assessment language (i.e., assessed in English, or another language), depending on sufficient studies.

## Results

To enhance organization of the Results section, we assigned a number to each paper included in the analyses. The corresponding numbers and papers are listed in S4. We refer to these numbers in the Results section as well as Tables 1-5 and Supplemental Materials S1-S18. All included papers are also cited across the current review. Supplemental Table S5 specifies extracted data items across studies, while Supplemental Excel File S6 indicates statistical values for associations between literacy precursors and reading outcomes as reported by reviewed studies.

**Table 1.**
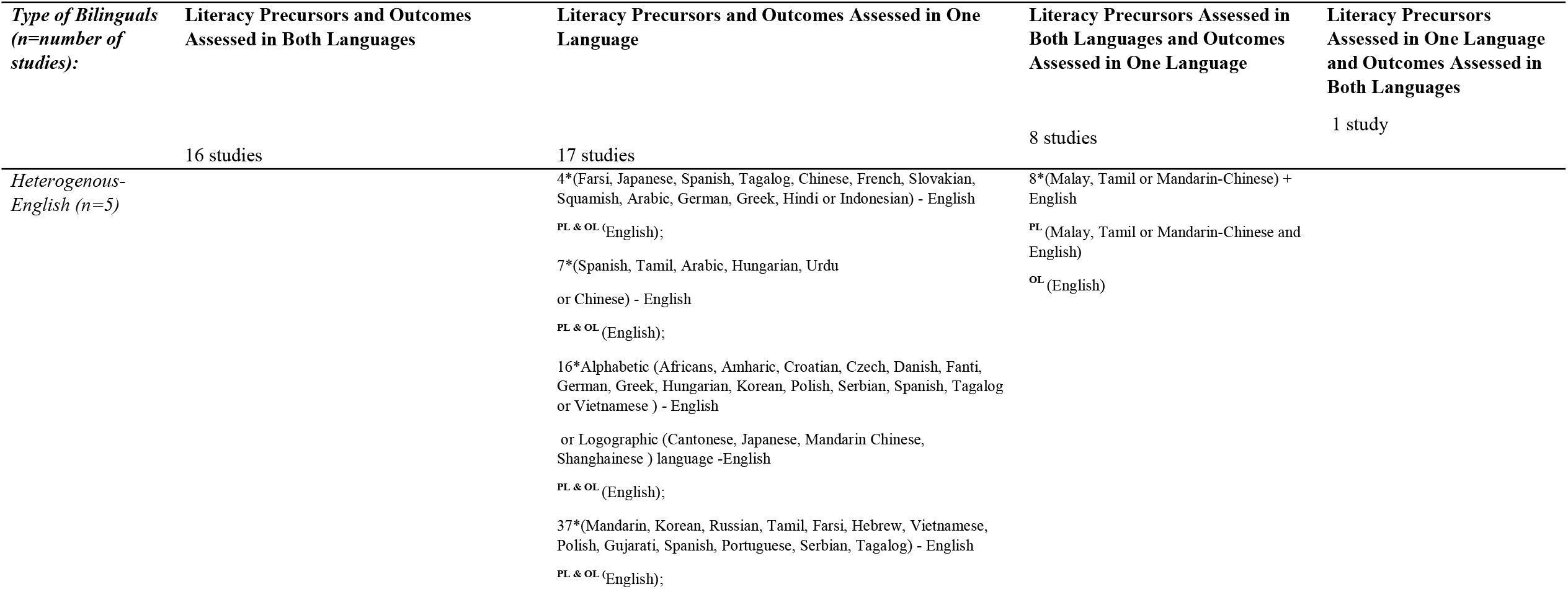
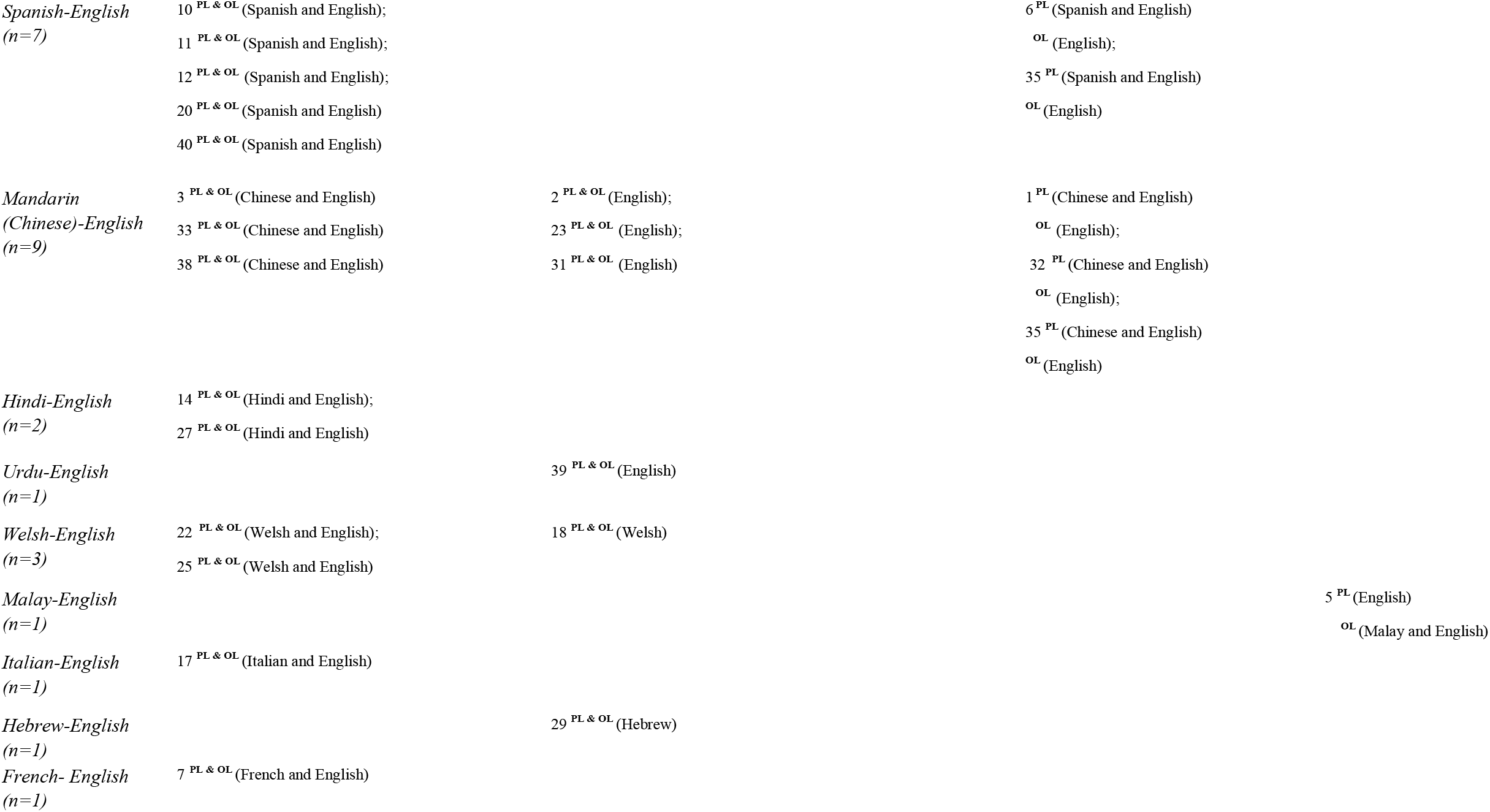
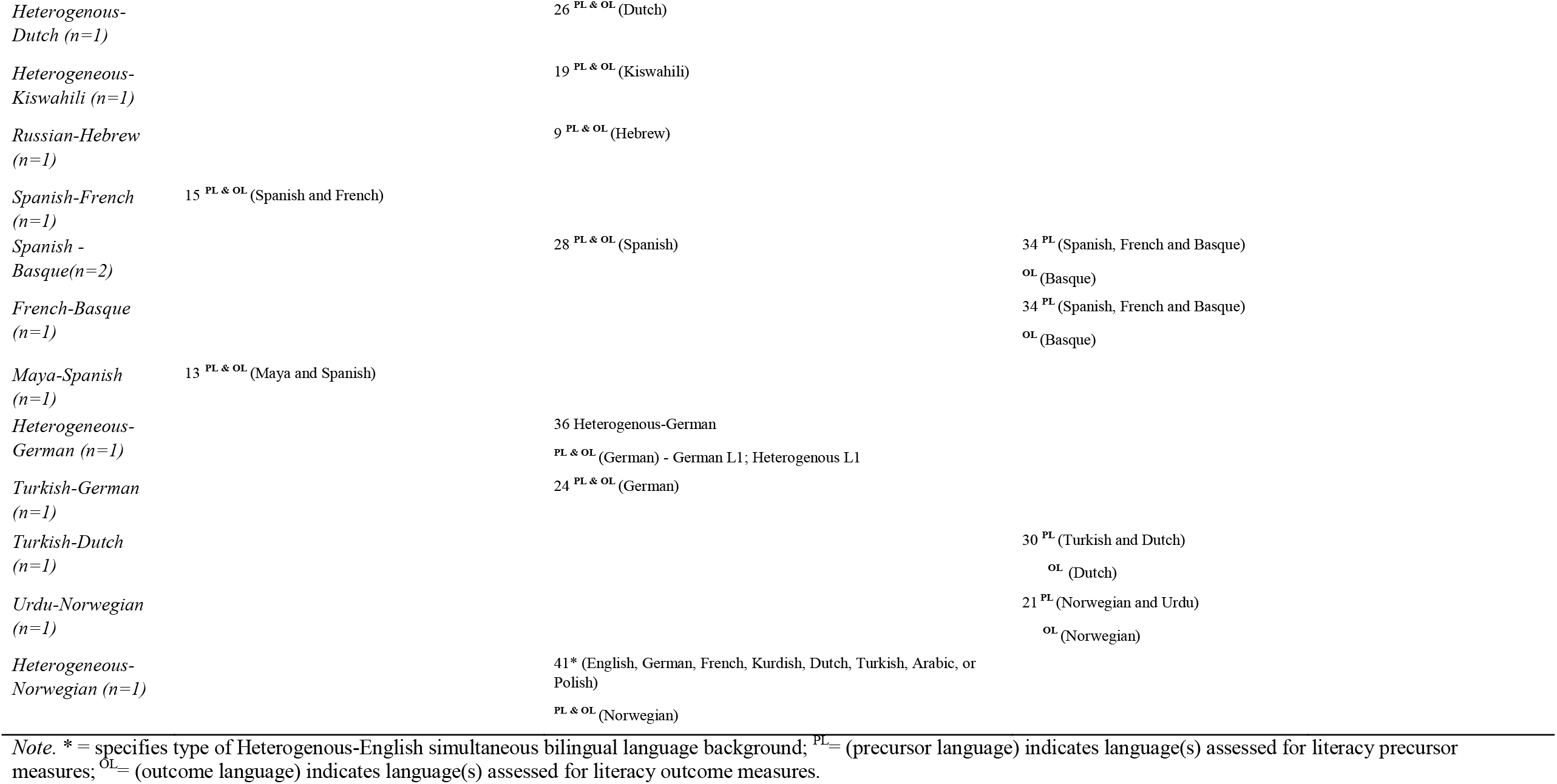
Literacy Precursors and Literacy Outcome Measures Assessed in One or Both Bilingual Languages, based on Bilingual Language Background. Table indicating the bilingual language background (i.e., combination of languages spoken and assessed, along with the number of studies: indicated on the left) of simultaneous bilinguals and the language(s) of literacy precursor and literacy outcome measure assessment (i.e., whether assessed in one or both bilingual languages) for each study.

### Database Search

After screening and assessment, 56 studies across 41 papers, comprising peer-reviewed (n=39) and gray literature (n=2; see S4 for further details) were included, with publication years ranging from 1977 – 2022. Our Figure 1 PRISMA Flow Diagram further illustrates this process (Page et al., 2021).

**Figure 1.**
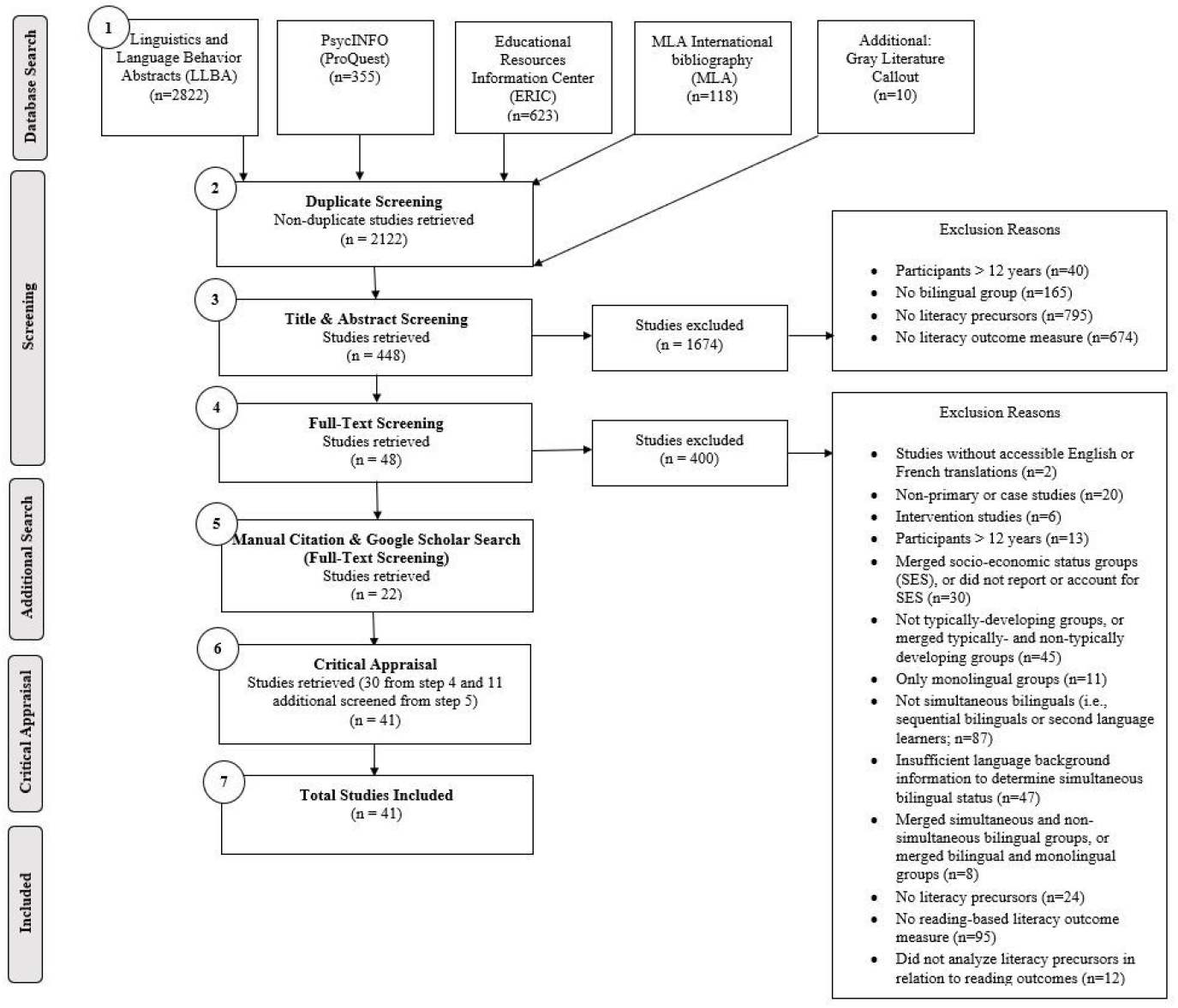
*Preferred Reporting Items for Systematic Reviews and Meta-Analyses (PRISMA) Flow Diagram for the Systematic Literature Review Search*. PRISMA Flow Diagram specifying number of studies included at title/abstract, full-text, additional Google Scholar and manual citation search, and critical appraisal levels of screening.

### Participants

The 41 reviewed studies represented children from diverse language backgrounds, speaking 21 bilingual language combinations. The most assessed groups were Chinese (Mandarin)-English (n=9), Spanish-English (n=7), and Heterogeneous-English (n=5) simultaneous bilinguals. The average age at assessment was 7;5 years. Participants were predominantly assessed in countries where English is the societal/educational language (n=17), including Canada (n=8) and the US (n=9). Other countries include Singapore, Spain (n=4), India, Wales (n=3), Netherlands, Germany, Norway (n=2), China, Kenya, Middle-East, and the UK (n=1). See S5 for age and country of assessment.

The reviewed papers assessed children who speak any combination of heritage and societal languages – across orthography type (i.e., alphabetic or non-alphabetic), orthographic depth (i.e., degree of consistency between symbol-sound correspondences - whether transparent or opaque), and linguistic distance (i.e., percentage of shared phonologically or semantically-related cognates across languages, and linguistic structures). Despite this relative variation in languages, literacy precursors and reading outcomes were predominantly assessed in English (n=28). Other assessed languages include Spanish (n=11), Chinese (n=7), Welsh, French (n=3), Hebrew, Hindi, Dutch, German, Norwegian (n=2), Turkish, Malay, Italian, Swahili, Maya, and Urdu (n=1). Table 1 specifies the language(s) of assessment for literacy precursor and outcome measures.

### Literacy Precursors Assessed in Relation to Word/Non-Word and Text Reading

The most commonly-assessed *code-related precursor skills* were phonological awareness (n=19: 1, 2, 3, 4, 7, 8, 9, 14, 15, 19, 22, 23, 24, 32, 33, 35, 36, 38, 40), followed by letter knowledge [n=2: 4, 13], and RAN [n=6: 4, 22, 26, 33, 36, 38]). In terms of *oral-language/grammar skills*, the following precursors were assessed: vocabulary (receptive: n=21: 1, 3, 8, 10, 11, 16, 19, 21, 22, 23, 25, 26, 30, 33, 35, 36, 37, 38, 39, 40, 41; expressive: n=5, 6, 24, 33, 38, 39), grammar (syntactic awareness: n=5, 4, 17, 33, 36, 37; morphological awareness: n=11:1, 3, 5, 23, 33, 35, 36, 37, 38, 40, 41), and oral-language comprehension (n=4: 10, 11, 16, 41). The *domain-general cognitive skills* included: working/verbal short-term memory (n=6: 4, 17, 31, 33,36, 39), and non-verbal intelligence (n=8: 3, 22, 31, 32, 33, 37, 38, 39). *Decoding* (n=10: 21, 23, 24, 26, 30, 33, 37, 39, 40, 41) was assessed in relation to reading comprehension.

Nine *additional literacy precursors* (that do not fit the categories above) were also identified by this review. These include semantic awareness (n=3:7, 9, 26), spelling (n=3: 4, 17, 38), visual attention (VA) span (n=2:15, 34), orthographic processing skills (n=2: 2, 38), environmental print awareness (study 4), name writing (13), sub-lexical/phonological speech perception (28), sentence priming (29), and novel word learning (39). See Supplemental Table S7 for precursor-outcome associations between all precursors and given word and text-level reading and comprehension outcome measures. Literacy precursors and reading outcome measures were assessed in either one or both languages of the bilingual children. Literacy precursors with the most consistent precursor-outcome associations were *code-related and oral-language skills*. See Supplemental Tables S8-S9 for further details across assessed languages. Another relevant pattern is that *semantic awareness* always emerged as an unreliable precursor (studies 7, 9, 26; see S7).

### Correlational Meta-Analyses for Code-Related, Oral-Language, and Decoding Precursors

As Table 2 below demonstrates, *phonological awareness, vocabulary*, and *morphological awareness* were significantly associated with word/non-word reading, when assessed in English and in another heritage or societal language. See Supplemental Figures S10-S14 for meta-analysis plots pertaining to phonological awareness (English: S10; Another language: S11), vocabulary (English: S12; Another language: S13), and morphological awareness (S14).

**Table 2.**
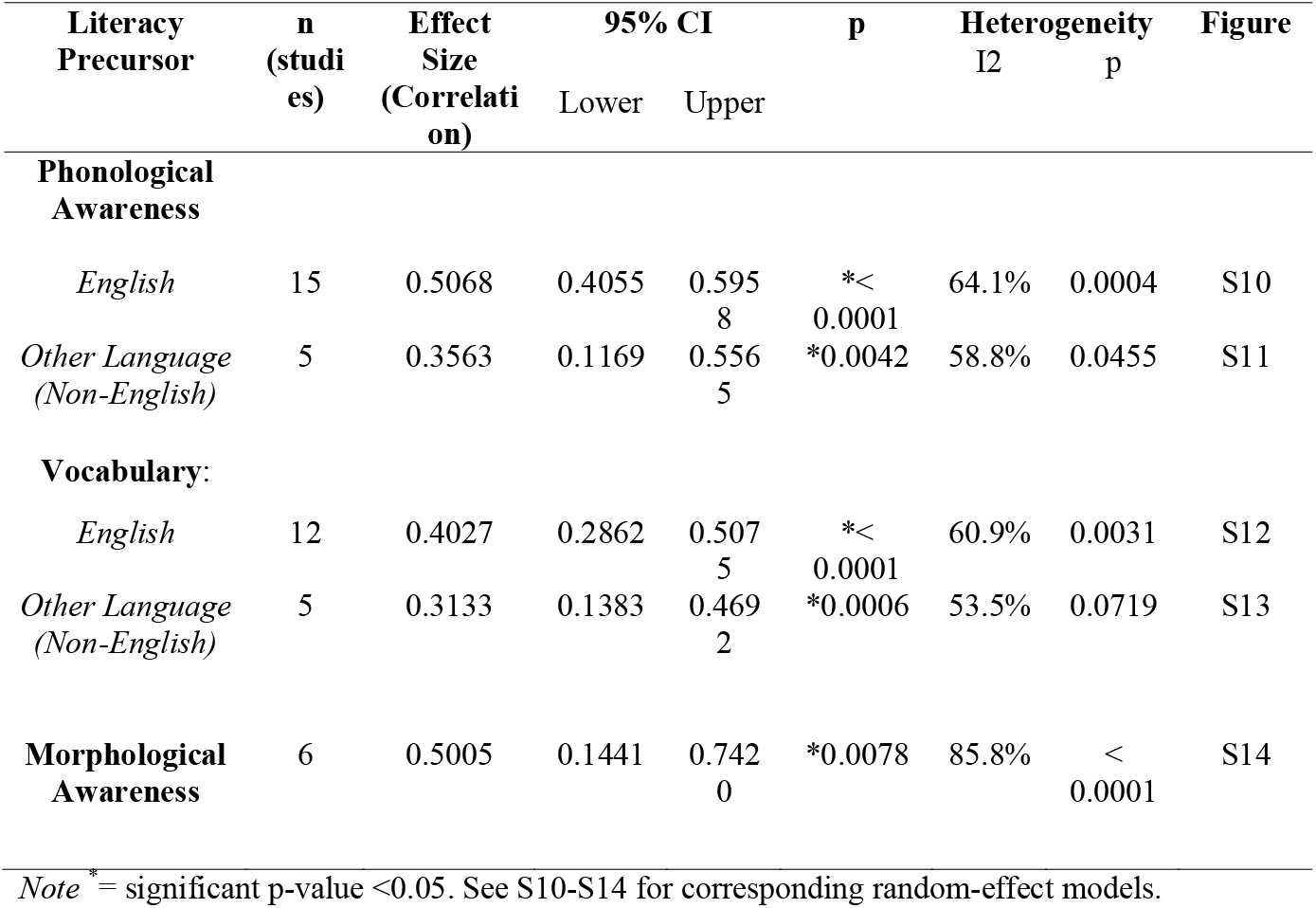
Correlational Meta-Analyses Results Across Code-Related and Oral-Language Precursor Type, in relation to Word and Non-Word Reading. We listed correlational effect size, along with number of studies (n), 95% CI, p-values and heterogeneity values, for **phonological awareness, vocabulary** and **morphological awareness**, in relation to word/non-word reading. When possible, we conducted two analyses, based on the assessment language (i.e., whether assessed in English only, or another language only).

As Table 3 below demonstrates, our analyses were significant for *vocabulary* and *word/non-word decoding* precursors in relation to text reading comprehension as well. See Supplemental Figures S15 and S16 for meta-analysis plots.

**Table 3.**
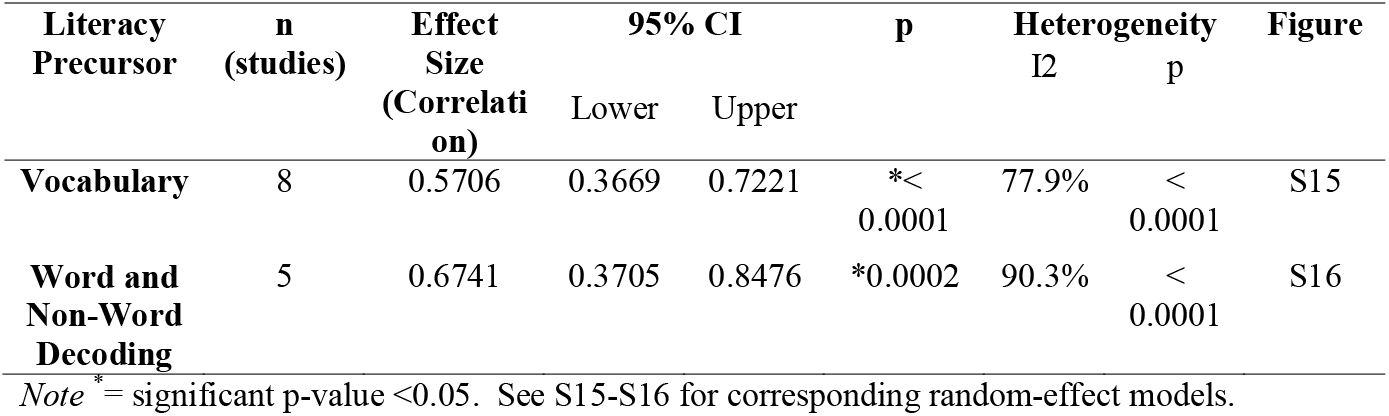
Correlational Meta-Analyses Results Across Precursor Type, in relation to Text Reading Comprehension. We listed correlational effect size, along with number of studies (n), 95% CI, p-values and heterogeneity values, for **vocabulary** and **word/non-word decoding**, in relation to text comprehension.

We conducted subgroup meta-analyses for phonological awareness in relation to word/non-word reading, based on whether the precursors and outcomes were assessed within the same language (e.g., English phonological awareness in relation to English word reading) or cross-language (e.g., English phonological awareness in relation to Welsh or Chinese word reading). As shown in Table 4 below, while the overall model was significant, *testing* condition (i.e., within versus cross-language) was not significant but close to significance. Supplemental Figure S17 also shows that within-language assessments had a larger effect size compared to cross-language assessments. Overall, it seems that within language assessments are more reliable than cross-language ones; however, it should be noted that we were only able to gather 5 data points for the cross-language group. This sample size limitation may have influenced Type-II error rate.

**Table 4.**
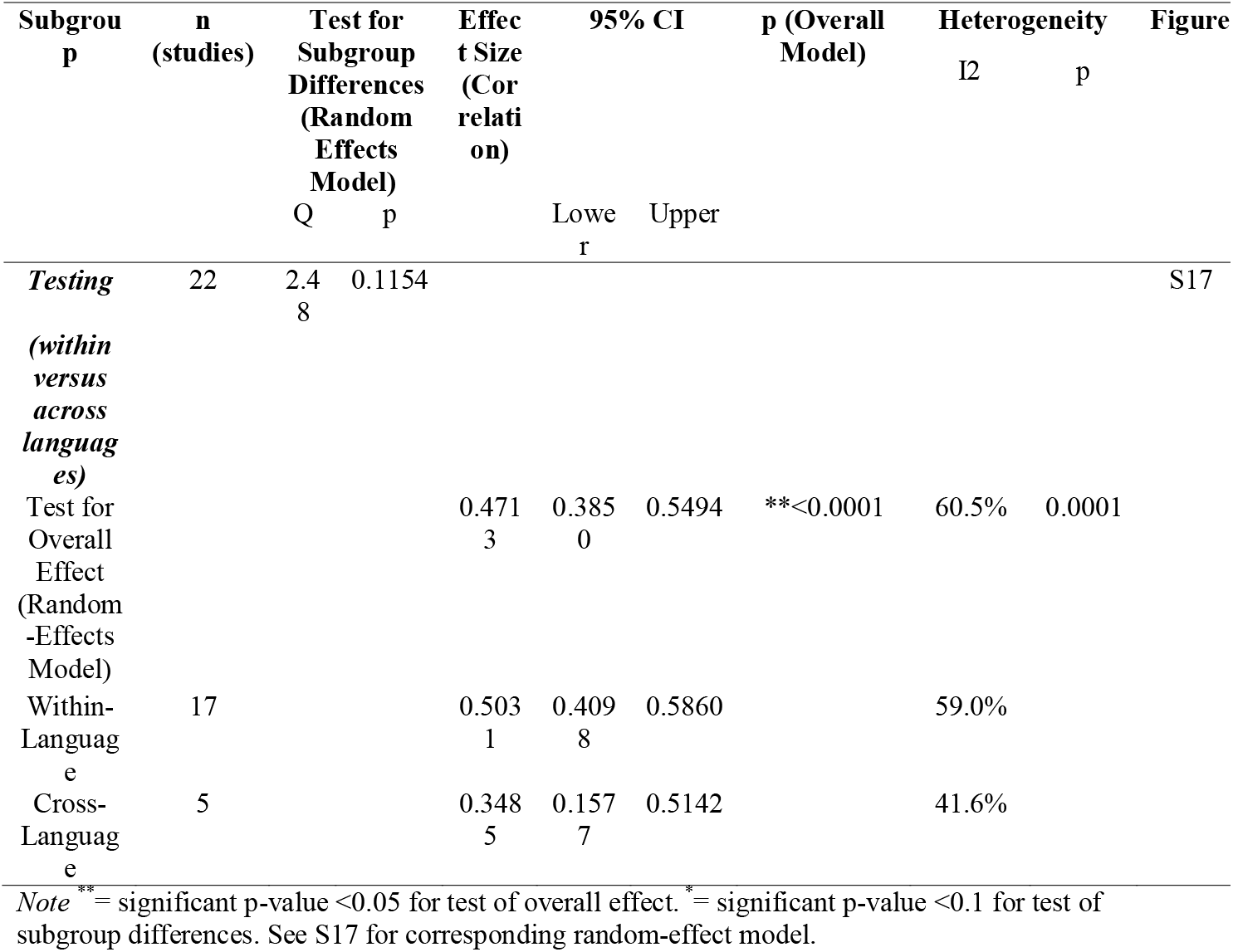
Subgroup Analyses for Phonological Awareness in relation to Word/Non-Word Reading, based on Type of Literacy Precursor and Outcome Measure Testing. We listed the test for overall correlational meta-analysis effect size, along with number of studies (n), test for subgroup differences, 95% CI, p-values and heterogeneity values, for **phonological awareness** in relation to word/non-word reading as an outcome measure, based on the type of *testing* subgroup (i.e., whether phonological awareness and word/non-word reading measures were assessed in the same language [within-language testing] or different languages [cross-language testing]).

When the same within versus cross-language comparison was considered for vocabulary as precursor, the overall model and subgroup tests were significant, as Table 5 demonstrates. This indicates that there are significant correlational differences based on whether the vocabulary precursor and word reading outcome measures were assessed within the same or different languages. Considering these results and the subgroup analysis plot presented in Supplemental Figure S18, it appears that within-language assessments are more reliable than cross-language ones.

**Table 5.**
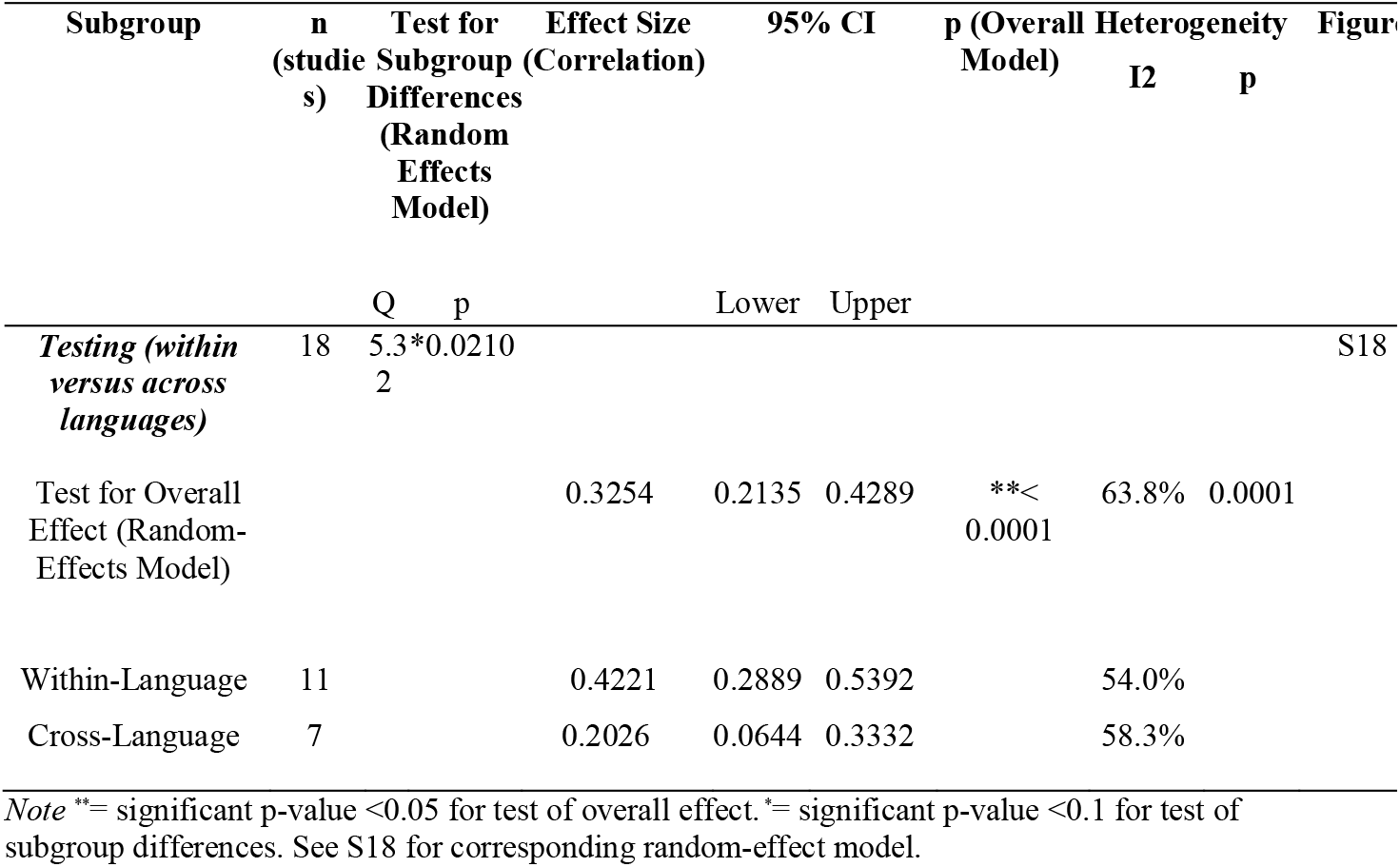
Subgroup Analyses for Vocabulary in relation to Word/Non-Word Reading, based on Type of Literacy Precursor and Outcome Measure Testing. We listed the test for overall correlational meta-analysis effect size, along with number of studies (n), test for subgroup differences, 95% CI, p-values and heterogeneity values, for **vocabulary** in relation to word/non-word reading as an outcome measure, based on the type of *testing* subgroup (i.e., whether vocabulary and word/non-word reading measures were assessed in the same language [within-language testing] or different languages [cross-language testing]).

## Discussion

This review synthesized findings from 56 studies across 41 papers. Overall, we found that the nine *code-related, oral-language, domain-general cognitive*, and *word/non-word decoding* precursors predict within- and cross-language reading outcomes in simultaneous bilingual children, across heritage and societal languages. In general, similar to English monolinguals (Hjetland et al., 2017, 2020; NELP, 2008), *code-related* (phonological awareness: Cherodath & Singh, 2015; Chiappe et al., 2002; Hipfner-Boucher et al., 2014; Hsu et al., 2016; Ibrahim et al., 2007; Jasińska & Petitto, 2017; Jasińska et al., 2019; Lallier et al., 2014; Lam et al., 2012; Limbird et al., 2014; Mak, 2013 ; Marks et al., 2022; Novita et al., 2022; O’Brien et al., 2019; Ruan et al., 2022; Spencer & Hanley, 2010; Sun et al., 2022; Yang, 2010; Yeong et al., 2014) and *oral-language* (receptive and expressive vocabulary: Babayiğit et al., 2022; Bérubé & Marinova-Todd, 2011; Dunn et al., 2011; Gunnerud et al., 2022; Hammer et al., 2007; Hipfner-Boucher et al., 2014; Hsu et al., 2016; Jasińska et al., 2019; Lam et al., 2012; Lervåg & Aukrust, 2010; Limbird et al., 2014; Mak, 2013 ; Marks et al., 2022; Novita et al., 2022; O’Brien et al., 2019; Peets et al., 2019; Rhys & Thomas, 2012; Ruan et al., 2022; Spatgens & Schoonen, 2017; Spencer & Hanley, 2010; Sun et al., 2022; Tamis-LeMonda et al., 2014; van den Bosch et al., 2020; Zhang, 2015) skills were commonly-assessed precursors.

In our meta-analyses, phonological awareness was reliably associated with word/non-word reading, and vocabulary was associated with reading comprehension (RC). Significant correlations were also evident between morphological awareness and word/non-word reading (Hipfner-Boucher et al., 2014; Hsu et al., 2016; Lam et al., 2012; Mak, 2013 ; see Table 2); as well as for vocabulary (Bérubé & Marinova-Todd, 2011; Lam et al., 2012; Limbird et al., 2014; Rhys & Thomas, 2012; van den Bosch et al., 2020; Mak, 2013) and word decoding (Lam et al., 2012; Limbird et al., 2014; van den Bosch et al., 2020; Mak, 2013; see Table 3) in relation to text comprehension. Our meta-analyses for phonological awareness and vocabulary measures also suggest that these precursors work best for assessment purposes when examined within the same language (e.g., English phonological awareness predicting English word reading; Hipfner-Boucher et al., 2014), as compared to when measured across languages (e.g., English phonological awareness predicting Welsh or Chinese word reading; Hipfner-Boucher et al., 2014; Spencer & Hanley, 2010; see Tables 4 and 5).

Many studies included here assessed bilinguals using standardized measures normed on English monolingual populations (also see summary in S5 for further details). These include the Comprehensive Test of Phonological Processing (CTOPP; Wagner at al., 2013; n=8/19) for phonological processing and Peabody Picture Vocabulary Test (PPVT; Dunn, 2019; n=12/21) for receptive vocabulary. When assessed in heritage languages related to English such as Spanish, these measures were based on the English standardized tests. For assessments in non-European languages such as Hebrew or Chinese, measures were often developed for research purposes and had not been previously used for literacy assessment in clinical practice for either monolinguals or bilinguals (Ibrahim et al., 2007; Ruan et al., 2022). These English-centric assessment practices reflect the lack of linguistic diversity in child development research, and further highlight the need for linguistically- and culturally-responsive literacy assessment tools to ensure comprehensive evaluation.

Despite this clear English- or monolingual-centric assessment limitation, our meta-analyses demonstrated a consistent relationship between emergent literacy precursor measures and various reading outcomes across a diverse bilingual population. In other words, these literacy precursors, typically assessed via tools developed for monolinguals, seem to work in bilinguals as well. However, two points should be considered when interpreting this finding. First, the most commonly-assessed literacy precursors in the current review were code-related skills (e.g., phonological awareness). It is a possibility that these sorts of meta-linguistic assessments are less affected by the child’s language background (i.e., monolingual versus bilingual); these precursors involve language-universal cognitive manipulation in addition to language-specific linguistic processes. Second, we examined simultaneous bilingual populations, thereby limiting age of acquisition (AoA) and language proficiency/dominance influences on assessment performance. Most of the children included here were proficient in both of their languages at the time of assessment. More than half of reviewed studies (n=28/41) assessed English-speaking simultaneous bilinguals, with an early AoA prior to 3 years, in English and/or another heritage or societal language such as Hindi (e.g., Gupta & Jamal, 2007). This may also explain the results for bilinguals when assessed on measures normed for English monolinguals. Clinicians and researchers should therefore not extend our findings to other types of bilingual populations. Future meta-analyses should examine whether these literacy precursors, and assessment tools, also perform well with other bilingual populations such as sequential bilinguals who might have less proficiency in the language of assessment at the time of evaluation.

Not surprisingly, reading models reflect the same pattern of monolingual- or English-centered frameworks as well. The Linguistic Interdependence Hypothesis (LIH) and Script Dependent Hypothesis (SDH) primarily examine monolingual readers of alphabetic and non-alphabetic languages, and cross-language literacy transfer effects in alphabetic-alphabetic bilinguals with shared Germanic or Romance families (Ellis & Hooper, 2002; Nation, 2019). These models do not address relationship strength differences evident for certain precursors and reading measures, across languages varying in orthographic depth.

We also identified nine other relatively under-investigated precursors used with bilingual children. These are: *semantic awareness* (Ibrahim et al., 2007; Jasińska & Petitto, 2017; Spatgens & Schoonen, 2017), s*pelling* (Chiappe et al., 2002; D’angiulli et al., 2002; Ruan et al., 2022), *visual attention (VA) span* (Lallier et al., 2014; Lallier et al., 2021), *orthographic processing* (Yeong et al., 2014; Ruan et al., 2022), *environmental print awareness* (Chiappe et al., 2002), *name writing* (Bengochea et al., 2015), *sub-lexical/phonological speech perception* (Ríos-López et al., 2017), *sentence priming* (Vital & Karniol, 2010), and *novel word learning* (Babayiğit et al., 2022). These precursors are also associated with reading abilities of monolingual children (e.g., Bosse & Valdois, 2009; Both-de Vries & Bus, 2010; Nation & Snowling, 1998; Vanvooren et al., 2017). However, further research evaluating their reliability in linguistically-diverse monolingual and bilingual populations is needed. For instance, orthographic processing skills could be relevant to bilinguals learning to read in languages with disparate orthographic features. For Chinese-English bilinguals, cross-language transfer of lexical orthographic strategies from the morpho-syllabic Chinese script may facilitate balanced use of dual – compared to predominant sub-lexical – reading strategies in the alphabetic script (Yeong et al., 2014; Babayiğit et al., 2022).

Significant precursor-outcome associations were evident for eight of the nine precursors above, apart from *semantic awareness* (Ibrahim et al., 2007; Jasińska & Petitto, 2017; Spatgens & Schoonen, 2017). In reviewed studies that assessed both monolinguals and bilinguals, semantic awareness significantly predicted word and text-level reading for English, Hebrew and Arabic monolinguals – but not for age-matched bilinguals (Jasińska & Petitto, 2017; Ibrahim et al., 2007). The non-significant precursor-outcome associations were evident across different types of reading measure: word and text-level reading accuracy (Ibrahim et al., 2007; Jasińska & Petitto, 2017), fluency (Ibrahim et al., 2007) and comprehension (Spatgens & Schoonen, 2017). It is a possibility that the non-significant outcomes for semantic awareness may be due to comparative language dominance differences. The bilinguals were only assessed in the school language (Ibrahim et al., 2007; Jasińska & Petitto, 2017; Spatgens & Schoonen, 2017), which may not be the dominant language of spoken and written language. However, we could not examine whether language dominance mediates relationship strength between precursors and reading measures across assessed languages, due to limited reporting of language background factors in the reviewed papers. In addition, the construct validity of the semantic awareness measure may have influenced findings, as some studies assessed semantic knowledge/processing measures linked to vocabulary and categorical associations rather than higher-level semantic awareness skills (Ibrahim et al., 2017; Jasińska & Petitto, 2017; Spatgens & Schoonen, 2017). Bilingual children might need more time to develop these associations than monolinguals, depending on their proficiency in the assessed language.

Our systematic review and meta-analyses focused on simultaneous bilinguals, with an age of acquisition less than 3 years across both languages. While our review minimized heterogeneity by only focusing on one type of bilingual (i.e., simultaneous bilinguals), some other factors such as language proficiency and dominance remain unexamined. This is because very few reviewed studies reported proficiency/exposure measures. Only 4/41 studies reported degree of language exposure (Hsu et al., 2016; Sun et al., 2021; Yang, 2010; Yeong et al., 2014) with 3/4 studies reporting language exposure details in both languages, while no studies reported degree of language usage. For instance, Yeong et al. (2014) demonstrate influences of language exposure and assessment type on precursor skills. Chinese-English simultaneous bilinguals, with greater English home exposure, demonstrated significantly higher scores on English receptive vocabulary and phonological awareness measures (for phoneme and syllable blending, not elision), compared to bilinguals with greater Chinese exposure. It was also surprising that only few studies conducted some analysis of or acknowledged the need for examining language proficiency (D’angiulli et al., 2002; Kovelman et al., 2015; Oller et al., 2007; Ríos-López et al., 2017; Sun et al., 2021; Yang, 2010).

Moreover, only one study investigated how interactions between linguistic factors, such as linguistic distance and orthographic depth – whether transparent or opaque – in addition to language dominance, mediate relationships between certain precursors and outcome measures across the two languages of bilinguals (Gunnerud et al., 2022). As compared to phonological awareness and decoding, vocabulary is strongly associated with RC in transparent orthographies at an earlier age as readers quickly establish decoding skills due to consistent grapheme-phoneme correspondences. School-aged readers, such as Norwegian bilinguals, can therefore allocate cognitive and linguistic resources towards comprehending text (Gunnerud et al., 2022).

Orthographic depth may also interact with the assessed language’s comparative dominance level (Gunnerund et al., 2022), to influence emergent biliteracy development including cross-language transfer effects. Theories of cross-language transfer, such as the LIH (Cummins, 1979) and its extension – the Interactive Transfer Framework (Chung et al., 2019, also emphasize the role of language dominance and orthographic depth for reading. For example, the LIH highlights the role of heritage-language proficiency and established precursor skills for facilitating emergent literacy skills and reading development in the societal language. However, these two factors – language dominance and orthographic depth – remain relatively under-investigated in biliteracy research and clinical literacy assessments. Comprehensive reporting and examination of language background factors would allow a better understanding of the relevance of cross-language transfer and reading models for bilingual populations across language background and dominance profiles.

We observed that literacy precursors predicted reading skills not only within languages, but across both language combinations as well. These cross-language transfer effects are in line with Cross-Language Transfer Theories (Chung et al., 2019) including Central Processing Hypothesis (e.g., Durgunoğlu, 2002). The theories indicate that meta-linguistic precursors (e.g., phonological awareness) are conducive to transfer across disparate languages, due to underlying domain-general cognitive skills. These bidirectional cross-linguistic transfer effects emphasize assessing and supporting emergent literacy development across both spoken languages (Kim & Piper, 2019), as certain skills can transfer from one language to another in bilingual children (Melby□Lervåg & Lervåg, 2011). Because of this, it is important for clinicians to consider linguistically-appropriate precursor assessment tools and reading measures, based on the language(s) of assessment’s phonological grain-size unit and orthographic properties. This will allow for comprehensive assessment of bilingual children, including examining whether evident reading difficulties are due to language-general cognitive deficits if present in both languages; or language-specific linguistic or proficiency differences if only present in the less-dominant language. To do so, future research should focus on developing linguistically- and culturally-responsive assessment tools. Linguistic diversity in child speech-language and literacy research will facilitate assessment accessibility and early intervention across heritage and societal languages of bilingual readers from under-represented linguistic communities.

### Limitations

Most research in this domain examines correlational analyses and literacy precursors in isolation (e.g., Hjetland et al., 2017; NELP, 2008). As such, reported *precursor-outcome associations* in our synthesis and meta-analyses do not imply a 1:1 predictive relationship. We acknowledge that primary study statistical limitations extend to this review. This limitation also affirms the need for different approaches to examining literacy precursors in future studies.

The review indicates a research bias for literacy-related knowledge production in English-dominant countries, such as Canada and the US. The bias for primary studies published in English and from high-income English-speaking countries limits generalizability for non-English dominant countries with differing language dominance profiles.

Also, few studies specified home and school literacy environments, universal pre-literacy assessment guidelines such as age and frequency of evaluation across countries, and reading instruction methods used in schools. Hence this information could not be part of our analysis.

### Future Research & Clinical Directions

- To address the English- and monolingual-centric assessment bias, future studies should examine reliability of non-linguistic precursors (e.g., environmental print awareness, working memory, etc.) when assessment tools are not available in both languages. Moreover, research should enhance linguistic diversity in research and clinical practice, by developing linguistically-appropriate literacy precursor assessment tools.
- Future studies should also report and analyze adequate and detailed language background measures across both languages to understand the contribution of linguistic factors (e.g., language balance/dominance, proficiency, etc.) to literacy development in bilinguals.
- To improve current models of reading, future studies should examine interactions between various precursors, along with individual and combined contributions to reading, in linguistically-diverse bilinguals across proficiency profiles. Such research would improve reading models, such as *Simple View of Reading* (*SVR), Dual Route Model of Reading, Linguistic Interdependence Hypothesis (LIH)* and *Script Dependent Hypothesis* (*SDH)*, that are predominantly based on monolingual alphabetic readers.

## Data Availability

All Tables and Figures are included in the pre-print, and supplemental files are available in the submission

## Acknowledgements

We thank Kai Ian Leung, for her help with screening papers, along with University of Toronto librarians, Chad Crichton, Erica Nekolaichuk, Glyneva Bradley-Ridout, and Nadia Muhe, for helping with the database and search terms selection and selecting appropriate analyses methods.

## Footnotes

1 According to the *Psycholinguistic Grain Size Theory* (Ziegler & Goswami, 2005), phonological grain sizes are the most accessible and consistently represented (onto orthographic units/symbols such as graphemes or characters) speech sound units in a language. These sound segments range from smaller phonological grain sizes such as phonemes (which are mapped onto graphemes in alphabetic languages) to larger syllables, onset-rimes and morphemes (mapped onto characters in morpho-syllabic/logographic and alpha-syllabic languages such as Chinese or Tamil, with larger grain sizes). The consistency of the relationship between phoneme grain size units and corresponding orthographic units differs across alphabetic languages, based on orthographic depth. Orthographically-transparent systems demonstrate one-to-one grapheme-phoneme correspondences, in comparison to opaque systems.

2 We also verified full texts of abstracts on second and dual language learners (i.e., those children who learn the second language [L2] at school after the age of 6 in a non-L2 majority country), given that these terms are sometimes used interchangeably with the term simultaneous bilinguals.

