## Supplementary material for "Early Precursors of Literacy Development in Simultaneous Bilinguals: A Systematic Review and Meta-Analysis": Figure 1

*Preferred Reporting Items for Systematic Reviews and Meta-Analyses (PRISMA) Flow Diagram for the Systematic Literature Review Search.* PRISMA Flow Diagram specifying number of studies included at title/abstract, full-text, additional Google Scholar and manual citation search, and critical appraisal levels of screening.

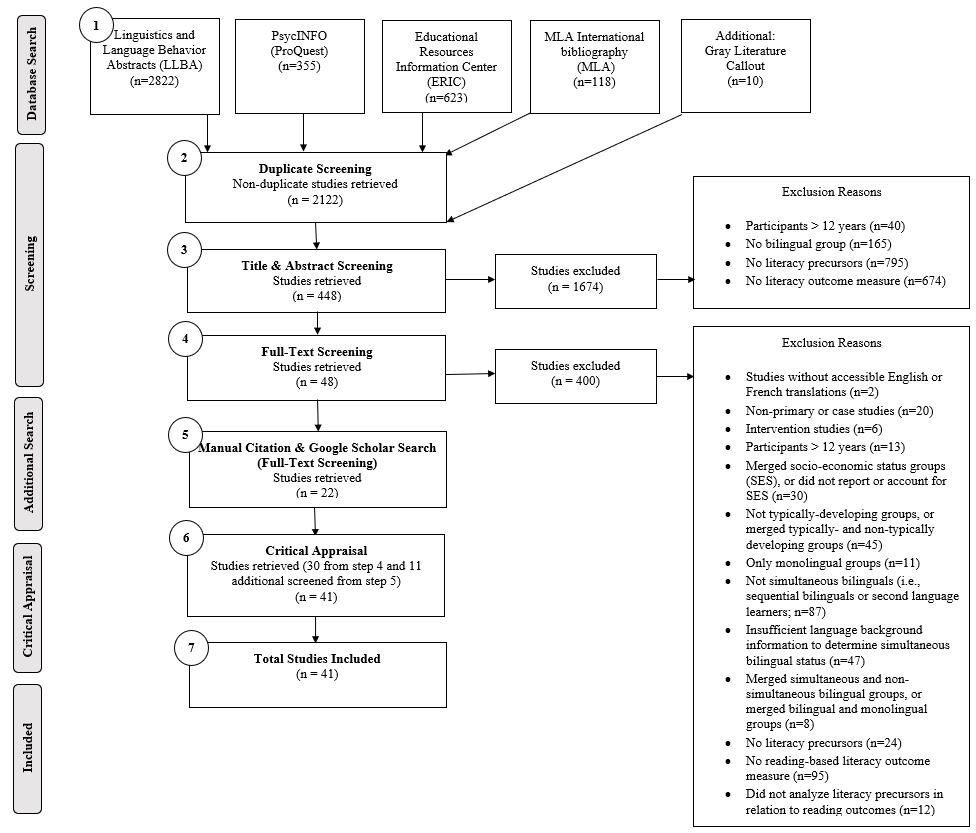
