## Supplementary material for "Early Precursors of Literacy Development in Simultaneous Bilinguals: A Systematic Review and Meta-Analysis": Table 1

*Literacy Precursors and Literacy Outcome Measures Assessed in One or Both Bilingual Languages, based on Bilingual Language Background.* Table indicating the bilingual language background (i.e., combination of languages spoken and assessed, along with the number of studies: indicated on the left) of simultaneous bilinguals and the language(s) of literacy precursor and literacy outcome measure assessment (i.e., whether assessed in one or both bilingual languages) for each study.

| ***Type of Bilinguals (n=number of studies):*** | **Literacy Precursors and Outcomes Assessed in Both Languages**      16 studies | **Literacy Precursors and Outcomes Assessed in One Language**      17 studies | **Literacy Precursors Assessed in Both Languages and Outcomes Assessed in One Language**    8 studies | **Literacy Precursors Assessed in One Language and Outcomes Assessed in Both Languages**    1 study |
| --- | --- | --- | --- | --- |
| *Heterogenous-English (n=5)* |  | 4*(Farsi, Japanese, Spanish, Tagalog, Chinese, French, Slovakian, Squamish, Arabic, German, Greek, Hindi or Indonesian) - English  **^PL & OL (^**English);  7*(Spanish, Tamil, Arabic, Hungarian, Urdu  or Chinese) - English  **^PL & OL^** (English);  16*Alphabetic (Africans, Amharic, Croatian, Czech, Danish, Fanti, German, Greek, Hungarian, Korean, Polish, Serbian, Spanish, Tagalog or Vietnamese ) - English  or Logographic (Cantonese, Japanese, Mandarin Chinese, Shanghainese ) language -English  **^PL & OL^** (English);  37*(Mandarin, Korean, Russian, Tamil, Farsi, Hebrew, Vietnamese, Polish, Gujarati, Spanish, Portuguese, Serbian, Tagalog) - English  **^PL & OL (^**English); | 8*(Malay, Tamil or Mandarin-Chinese) + English  **^PL^** (Malay, Tamil or Mandarin-Chinese and English)  **^OL^** (English) |  |
| *Spanish-English (n=7)* | 10 **^PL & OL^** (Spanish and English);  11 **^PL & OL^** (Spanish and English);  12 **^PL & OL^** (Spanish and English);  20 **^PL & OL^** (Spanish and English)  40 **^PL & OL^** (Spanish and English) |  | 6 **^PL^** (Spanish and English)  **^OL^** (English);  35 **^PL^** (Spanish and English)  **^OL^** (English) |  |
| *Mandarin (Chinese)-English (n=9)* | 3 **^PL & OL^** (Chinese and English)  33 **^PL & OL^** (Chinese and English)  38 **^PL & OL^** (Chinese and English) | 2 **^PL & OL^** (English);  23 **^PL & OL^** (English);  31 **^PL & OL^** (English) | 1 **^PL^** (Chinese and English)  **^OL^** (English);  32 **^PL^** (Chinese and English)  **^OL^** (English);  35 **^PL^** (Chinese and English)  **^OL^** (English) |  |
| *Hindi-English (n=2)* | 14 **^PL & OL^** (Hindi and English);  27 **^PL & OL^** (Hindi and English) |  |  |  |
| *Urdu-English (n=1)* |  | 39 **^PL & OL^** (English) |  |  |
| *Welsh-English (n=3)* | 22 **^PL & OL^** (Welsh and English);  25 **^PL & OL^** (Welsh and English) | 18 **^PL & OL^** (Welsh) |  |  |
| *Malay-English (n=1)* |  |  |  | 5 **^PL^** (English)  **^OL^** (Malay and English) |
| *Italian-English (n=1)* | 17 **^PL & OL^** (Italian and English) |  |  |  |
| *Hebrew-English (n=1)* |  | 29 **^PL & OL^** (Hebrew) |  |  |
| *French- English (n=1)* | 7 **^PL & OL^** (French and English) |  |  |  |
| *Heterogenous-Dutch (n=1)* |  | 26 **^PL & OL^** (Dutch) |  |  |
| *Heterogeneous-Kiswahili (n=1)* |  | 19 **^PL & OL^** (Kiswahili) |  |  |
| *Russian-Hebrew (n=1)* |  | 9 **^PL & OL^** (Hebrew) |  |  |
| *Spanish-French (n=1)* | 15 **^PL & OL^** (Spanish and French) |  |  |  |
| *Spanish - Basque(n=2)* |  | 28 **^PL & OL^** (Spanish) | 34 **^PL^** (Spanish, French and Basque)  **^OL^** (Basque) |  |
| *French-Basque (n=1)* |  |  | 34 **^PL^** (Spanish, French and Basque)  **^OL^** (Basque) |  |
| *Maya-Spanish (n=1)* | 13 **^PL & OL^** (Maya and Spanish) |  |  |  |
| *Heterogeneous-German (n=1)* |  | 36 Heterogenous-German  **^PL & OL^** (German) - German L1; Heterogenous L1 |  |  |
| *Turkish-German (n=1)* |  | 24 **^PL & OL^** (German) |  |  |
| *Turkish-Dutch (n=1)* |  |  | 30 **^PL^** (Turkish and Dutch)  **^OL^** (Dutch) |  |
| *Urdu-Norwegian (n=1)* |  |  | 21 **^PL^** (Norwegian and Urdu)  **^OL^** (Norwegian) |  |
| *Heterogeneous-Norwegian (n=1)* |  | 41* (English, German, French, Kurdish, Dutch, Turkish, Arabic, or Polish)  **^PL & OL^** (Norwegian) |  |  |

*Note.* * = specifies type of Heterogenous-English simultaneous bilingual language background; ^PL^= (precursor language) indicates language(s) assessed for literacy precursor measures; ^OL^= (outcome language) indicates language(s) assessed for literacy outcome measures.
