## Supplementary material for "Early Precursors of Literacy Development in Simultaneous Bilinguals: A Systematic Review and Meta-Analysis": Table 2

*Correlational Meta-Analyses Results Across Code-Related and Oral-Language Precursor Type, in relation to Word and Non-Word Reading.* We listed correlational effect size, along with number of studies (n), 95% CI, p-values and heterogeneity values, for **phonological awareness**, **vocabulary** and **morphological awareness**, in relation to word/non-word reading. When possible, we conducted two analyses, based on the assessment language (i.e., whether assessed in English only, or another language only).

| **Literacy Precursor** | **n (studies)** | **Effect Size (Correlation)** | **95% CI** | | **p** | **Heterogeneity** | | **Figure** |
| --- | --- | --- | --- | --- | --- | --- | --- | --- |
|  |  |  |  |  |  | I2 | p |  |
|  |  |  | Lower | Upper |  |  |  |  |
| **Phonological Awareness** |  |  |  |  |  |  |  |  |
| *English* | 15 | 0.5068 | 0.4055 | 0.5958 | *< 0.0001 | 64.1% | 0.0004 | S10 |
| *Other Language (Non-English)* | 5 | 0.3563 | 0.1169 | 0.5565 | *0.0042 | 58.8% | 0.0455 | S11 |
| **Vocabulary**: |  |  |  |  |  |  |  |  |
| *English* | 12 | 0.4027 | 0.2862 | 0.5075 | *< 0.0001 | 60.9% | 0.0031 | S12 |
| *Other Language (Non-English)* | 5 | 0.3133 | 0.1383 | 0.4692 | *0.0006 | 53.5% | 0.0719 | S13 |
| **Morphological Awareness** | 6 | 0.5005 | 0.1441 | 0.7420 | *0.0078 | 85.8% | < 0.0001 | S14 |

*Note* ^*^= significant p-value <0.05. See S10-S14 for corresponding random-effect models.
