## Supplementary material for "Early Precursors of Literacy Development in Simultaneous Bilinguals: A Systematic Review and Meta-Analysis": Table 3

*Correlational Meta-Analyses Results Across Precursor Type, in relation to Text Reading Comprehension.* We listed correlational effect size, along with number of studies (n), 95% CI, p-values and heterogeneity values, for **vocabulary** and **word/non-word decoding**, in relation to text comprehension.

| **Literacy Precursor** | **n (studies)** | **Effect Size (Correlation)** | **95% CI** | | **p** | **Heterogeneity** | | **Figure** |
| --- | --- | --- | --- | --- | --- | --- | --- | --- |
|  |  |  |  |  |  | I2 | p |  |
|  |  |  | Lower | Upper |  |  |  |  |
| **Vocabulary** | 8 | 0.5706 | 0.3669 | 0.7221 | *< 0.0001 | 77.9% | < 0.0001 | S15 |
| **Word and Non-Word Decoding** | 5 | 0.6741 | 0.3705 | 0.8476 | *0.0002 | 90.3% | < 0.0001 | S16 |

*Note* ^*^= significant p-value <0.05. See S15-S16 for corresponding random-effect models.
