## Supplementary material for "Early Precursors of Literacy Development in Simultaneous Bilinguals: A Systematic Review and Meta-Analysis": Table 4

*Subgroup Analyses for Phonological Awareness in relation to Word/Non-Word Reading, based on Type of Literacy Precursor and Outcome Measure Testing.* We listed the test for overall correlational meta-analysis effect size, along with number of studies (n), test for subgroup differences, 95% CI, p-values and heterogeneity values, for **phonological awareness** in relation to word/non-word reading as an outcome measure, based on the type of *testing* subgroup (i.e., whether phonological awareness and word/non-word reading measures were assessed in the same language [within-language testing] or different languages [cross-language testing]).

| **Subgroup** | **n (studies)** | **Test for Subgroup Differences (Random Effects Model)** | | **Effect Size (Correlation)** | **95% CI** | | | **p (Overall Model)** | | **Heterogeneity** | | | | **Figure** | |
| --- | --- | --- | --- | --- | --- | --- | --- | --- | --- | --- | --- | --- | --- | --- | --- |
|  |  |  |  |  |  |  |  |  |  | I2 | | p | |  | |
|  |  | Q | p |  | | Lower | Upper | |  | |  | |  | |  |
| ***Testing***  ***(within versus across languages)*** | 22 | 2.48 | 0.1154 |  | |  |  | |  | |  | |  | | S17 |
| Test for Overall Effect (Random-Effects Model) |  |  |  | 0.4713 | | 0.3850 | 0.5494 | | **<0.0001 | | 60.5% | | 0.0001 | |  |
| Within-Language | 17 |  |  | 0.5031 | | 0.4098 | 0.5860 | |  | | 59.0% | |  | |  |
| Cross-Language | 5 |  |  | 0.3485 | | 0.1577 | 0.5142 | |  | | 41.6% | |  | |  |

*Note* ^**^= significant p-value <0.05 for test of overall effect. ^*^= significant p-value <0.1 for test of subgroup differences. See S17 for corresponding random-effect model.
