## Supplementary material for "Early Precursors of Literacy Development in Simultaneous Bilinguals: A Systematic Review and Meta-Analysis": Table 5

*Subgroup Analyses for Vocabulary in relation to Word/Non-Word Reading, based on Type of Literacy Precursor and Outcome Measure Testing.* We listed the test for overall correlational meta-analysis effect size, along with number of studies (n), test for subgroup differences, 95% CI, p-values and heterogeneity values, for **vocabulary** in relation to word/non-word reading as an outcome measure, based on the type of *testing* subgroup (i.e., whether vocabulary and word/non-word reading measures were assessed in the same language [within-language testing] or different languages [cross-language testing]).

| **Subgroup** | **n (studies)** | **Test for Subgroup Differences (Random Effects Model)** | | **Effect Size (Correlation)** | **95% CI** | | **p (Overall Model)** | **Heterogeneity** | | **Figure** |
| --- | --- | --- | --- | --- | --- | --- | --- | --- | --- | --- |
|  |  |  |  |  |  |  |  | **I2** | **p** |  |
|  |  | Q | p |  | Lower | Upper |  |  |  |  |
| ***Testing (within versus across languages)*** | 18 | 5.32 | *0.0210 |  |  |  |  |  |  | S18 |
| Test for Overall Effect (Random-Effects Model) |  |  |  | 0.3254 | 0.2135 | 0.4289 | **< 0.0001 | 63.8% | 0.0001 |  |
| Within-Language | 11 |  |  | 0.4221 | 0.2889 | 0.5392 |  | 54.0% |  |  |
| Cross-Language | 7 |  |  | 0.2026 | 0.0644 | 0.3332 |  | 58.3% |  |  |

*Note* ^**^= significant p-value <0.05 for test of overall effect. ^*^= significant p-value <0.1 for test of subgroup differences. See S18 for corresponding random-effect model.
