## Supplemental Table 1 for "Early Precursors of Literacy Development in Simultaneous Bilinguals: A Systematic Review and Meta-Analysis"

**S1**

*Full Electronic Database Search Strategy for the Four Databases.* Table of the full electronic search strategy, along with the search fields and limits specified, for the four Linguistics and Language Behavior Abstracts (LLBA), Educational Resources Information Center (ERIC), MLA International Bibliography (MLA) and PsycINFO (ProQuest) databases.

| **Database Search Terms** | | | | **Search Fields** | **Search Limits** |
| --- | --- | --- | --- | --- | --- |
| Linguistics and Language Behavior Abstracts (LLBA) | Educational Resources Information Center (ERIC) | MLA International Bibliography (MLA) | PsycINFO (ProQuest) |  |  |
| (Phonological Awareness *OR* phonem* awareness *OR* letter knowledge *OR* grapheme knowledge *OR* Grapheme Phoneme Correspondence *OR* sound symbol* *OR* Serial Recall *OR* Oral Comprehension *OR* Verbal Comprehension *OR* Listening Comprehension *OR* Vocabulary *OR* Word Knowledge *OR* grammar *OR* Syntax *OR* Syntac* *OR* Synta* *OR* Morpholog* *OR* Morphem* *OR* Visual Short Term Memory *OR* Phonological Short Term Memory *OR* Short Term Memory *OR* Working Memory *OR* Visual Memory *OR* Verbal Memory OR Nonverbal Memory *OR* Nonverbal Ability *OR* Nonverbal Intelligence *OR* Nonverbal IQ *OR* Precursor Literacy *OR* Precursor Reading *OR* Predictor Literacy *OR* Predictor Reading *OR* Precursor Literacy Skills *OR* Precursors of Reading Ability *OR* Early Predictors of Later Conventional Literacy Skills *OR* Predictive Literacy Skills *OR* Predictors of Later Reading Skills *OR* Preschool Literacy) *AND* (Word Decoding *OR* Reading Fluency *OR* Word Recognition *OR* Reading Ability *OR* Reading Skills *OR* Literacy Skills *OR* Reading Comprehension) *AND* mainsubject (Child* *OR* Infants) *AND* mainsubject (Bilingual* *OR* Multilingual* *OR* Second Language Learner*) | (Phonological Awareness *OR* phonem* awareness *OR* letter knowledge *OR* grapheme knowledge *OR* Grapheme Phoneme Correspondence *OR* sound symbol* *OR* Serial Recall *OR* Oral Comprehension *OR* Verbal Comprehension *OR* Listening Comprehension *OR* Vocabulary *OR* Word Knowledge *OR* grammar *OR* Syntax *OR* Syntac* *OR* Synta* *OR* Morpholog* *OR* Morphem* *OR* Visual Short Term Memory *OR* Phonological Short Term Memory *OR* Short Term Memory *OR* Working Memory *OR* Visual Memory *OR* Verbal Memory OR Nonverbal Memory *OR* Nonverbal Ability *OR* Nonverbal Intelligence *OR* Nonverbal IQ *OR* Precursor Literacy *OR* Precursor Reading *OR* Predictor Literacy *OR* Predictor Reading *OR* Precursor Literacy Skills *OR* Precursors of Reading Ability *OR* Early Predictors of Later Conventional Literacy Skills *OR* Predictive Literacy Skills *OR* Predictors of Later Reading Skills *OR* Preschool Literacy) *AND* (Word Decoding *OR* Reading Fluency *OR* Word Recognition *OR* Reading Ability *OR* Reading Skills *OR* Literacy Skills *OR* Reading Comprehension) *AND* mainsubject (Child* *OR* Infants) *AND* mainsubject (Bilingual* *OR* Multilingual* *OR* Second Language Learner*) | (Phonological Awareness *OR* phonem* awareness *OR* letter knowledge *OR* grapheme knowledge *OR* Grapheme Phoneme Correspondence *OR* sound symbol* *OR* Serial Recall *OR* Oral Comprehension *OR* Verbal Comprehension *OR* Listening Comprehension *OR* Vocabulary *OR* Word Knowledge *OR* grammar *OR* Syntax *OR* Syntac* *OR* Synta* *OR* Morpholog* *OR* Morphem* *OR* Visual Short Term Memory *OR* Phonological Short Term Memory *OR* Short Term Memory *OR* Working Memory *OR* Visual Memory *OR* Verbal Memory OR Nonverbal Memory *OR* Nonverbal Ability *OR* Nonverbal Intelligence *OR* Nonverbal IQ *OR* Precursor Literacy *OR* Precursor Reading *OR* Predictor Literacy *OR* Predictor Reading *OR* Precursor Literacy Skills *OR* Precursors of Reading Ability *OR* Early Predictors of Later Conventional Literacy Skills *OR* Predictive Literacy Skills *OR* Predictors of Later Reading Skills *OR* Preschool Literacy) *AND* (Word Decoding *OR* Reading Fluency *OR* Word Recognition *OR* Reading Ability *OR* Reading Skills *OR* Literacy Skills *OR* Reading Comprehension) *AND* mainsubject (Child* *OR* Infants) *AND* mainsubject (Bilingual* *OR* Multilingual* *OR* Second Language Learner*) | (Phonological Awareness *OR* phonem* awareness *OR* letter knowledge *OR* grapheme knowledge *OR* Grapheme Phoneme Correspondence *OR* sound symbol* *OR* Serial Recall *OR* Oral Comprehension *OR* Verbal Comprehension *OR* Listening Comprehension *OR* Vocabulary *OR* Word Knowledge *OR* grammar *OR* Syntax *OR* Syntac* *OR* Synta* *OR* Morpholog* *OR* Morphem* *OR* Visual Short Term Memory *OR* Phonological Short Term Memory *OR* Short Term Memory *OR* Working Memory *OR* Visual Memory *OR* Verbal Memory OR Nonverbal Memory *OR* Nonverbal Ability *OR* Nonverbal Intelligence *OR* Nonverbal IQ *OR* Precursor Literacy *OR* Precursor Reading *OR* Predictor Literacy *OR* Predictor Reading *OR* Precursor Literacy Skills *OR* Precursors of Reading Ability *OR* Early Predictors of Later Conventional Literacy Skills *OR* Predictive Literacy Skills *OR* Predictors of Later Reading Skills *OR* Preschool Literacy) *AND* (Word Decoding *OR* Reading Fluency *OR* Word Recognition *OR* Reading Ability *OR* Reading Skills *OR* Literacy Skills *OR* Reading Comprehension) *AND* mainsubject (Child* *OR* Infants) *AND* mainsubject (Bilingual* *OR* Multilingual* *OR* Second Language Learner*) | All | None |

*Note. AND* = mandatory search terms; *OR* = optional search terms; *** = truncation symbol to retrieve multiple search terms with a common root in the Linguistics and Language Behavior Abstracts, Educational Resources Information Center, MLA International Bibliography and PsycINFO (ProQuest) electronic databases.
