## Supplemental Table 2 for "Early Precursors of Literacy Development in Simultaneous Bilinguals: A Systematic Review and Meta-Analysis"

**S2: General Database Search Syntax**

Note:  For purposes of our database search, ‘*OR*’ is used to specify terms that are optional or alternatives for a specified term while ‘*AND*’ specifies terms that are mandatory.

***Database Search of the 8 Code-Related, Oral-Language, and Domain-General Cognitive* *Literacy Precursors (in Relation to Word/Non-Word Decoding or Reading)***

(Phonological Awareness *OR* phonem* awareness *OR* letter knowledge *OR* grapheme knowledge *OR* Grapheme Phoneme Correspondence *OR* sound symbol* *OR* Serial Recall *OR* Oral Comprehension *OR* Verbal Comprehension *OR* Listening Comprehension *OR* Vocabulary *OR* Word Knowledge *OR* grammar *OR* Syntax *OR* Syntac* *OR* Synta* *OR* Morpholog* *OR* Morphem* *OR* Visual Short Term Memory *OR* Phonological Short Term Memory *OR* Short Term Memory *OR* Working Memory *OR* Visual Memory *OR* Verbal Memory *OR* Nonverbal Memory *OR* Nonverbal Ability *OR* Nonverbal Intelligence *OR* Nonverbal IQ) *AND* (Word Decoding *OR* Reading Fluency *OR* Word Recognition *OR* Reading Ability *OR* Reading Skills *OR* Literacy Skills) *AND* (mainsubject(Child*) *OR* mainsubject(Infants)) *AND* (mainsubject(Bilingual*) *OR* mainsubject(Multilingual*) *OR* mainsubject(Second Language Learner*))

***Database Search of the 9 Code-Related, Oral-Language, and Domain-General Cognitive Literacy Precursors (in Relation to Reading Comprehension)***

(Phonological Awareness *OR* phonem* awareness *OR* letter knowledge *OR* grapheme knowledge *OR* Grapheme Phoneme Correspondence *OR* sound symbol* *OR* Serial Recall *OR* Oral Comprehension *OR* Verbal Comprehension *OR* Listening Comprehension *OR* Vocabulary *OR* Word Knowledge *OR* grammar *OR* Syntax *OR* Syntac* *OR* Synta* *OR* Morpholog* *OR* Morphem* *OR* Visual Short Term Memory *OR* Phonological Short Term Memory *OR* Short Term Memory *OR* Working Memory *OR* Visual Memory *OR* Verbal Memory *OR* Nonverbal Memory *OR* Nonverbal Ability *OR* Nonverbal Intelligence *OR* Nonverbal IQ  *OR* Word Decoding *OR* Reading Fluency *OR* Word Recognition *OR* Reading Ability *OR* Reading Skills OR Literacy Skills) *AND* (Reading Comprehension) *AND* (mainsubject(Child*) *OR* mainsubject(Infants)) *AND* (mainsubject(Bilingual*) *OR* mainsubject(Multilingual*) *OR* mainsubject(Second Language Learner*))

***Database Search of General/Additional Literacy Precursors (in Relation to Word/Non-Word Decoding or Reading)***

(Precursor Literacy *OR* Precursor Reading *OR* Predictor Literacy *OR* Predictor Reading *OR* Precursor Literacy Skills *OR* Precursors of Reading Ability *OR* Early Predictors of Later Conventional Literacy Skills *OR* Predictive Literacy Skills *OR* Predictors of Later Reading Skills *OR* Preschool Literacy) *AND* (Word Decoding *OR* Reading Fluency *OR* Word Recognition *OR* Reading Ability *OR* Reading Skills *OR* Literacy Skills) *AND* (mainsubject(Child*) *OR* mainsubject(Infants)) *AND* (mainsubject(Bilingual*) *OR* mainsubject(Multilingual*) *OR* mainsubject(Second Language Learner*))

***Database Search of General/Additional Literacy Precursors (in Relation to Reading Comprehension)***

(Precursor Literacy *OR* Precursor Reading *OR* Predictor Literacy *OR* Predictor Reading *OR* Precursor Literacy Skills *OR* Precursors of Reading Ability *OR* Early Predictors of Later Conventional Literacy Skills *OR* Predictive Literacy Skills *OR* Predictors of Later Reading Skills *OR* Preschool Literacy) *AND* (Reading Comprehension) *AND* (mainsubject(Child*) *OR* mainsubject(Infants)) *AND* (mainsubject(Bilingual*) *OR* mainsubject(Multilingual*) *OR* mainsubject(Second Language Learner*))
