## Supplemental File 3 for "Early Precursors of Literacy Development in Simultaneous Bilinguals: A Systematic Review and Meta-Analysis"

**S3**

*Database Search Terms for Literacy Precursors, Literacy Outcome Measures and Participant Demographic Characteristics.* Table of database search terms for the nine code-related, oral-language, and domain-general cognitive literacy precursors (phonological awareness, letter knowledge, RAN/serial recall, oral language/listening comprehension, vocabulary, grammar, memory, non-verbal intelligence and word decoding) and general/additional literacy precursors, two literacy outcome measures (word/non-word reading and text reading comprehension) and participant characteristics (age and type of language background) for the four Linguistics and Language Behavior Abstracts (LLBA), Educational Resources Information Center (ERIC), MLA International Bibliography (MLA) and PsycINFO (ProQuest) databases.

| **Literacy**  **Precursors** | **Phonological Awareness** | **Letter Knowledge** | **RAN/**  **Serial Recall** | **Oral Language/Listening Comprehension** | **Vocabulary** | **Grammar** | **Memory** | **Non-Verbal Intelligence** | **Word Decoding** | **General Literacy**  **Precursors** |
| --- | --- | --- | --- | --- | --- | --- | --- | --- | --- | --- |
| **Search Query** | Phonological Awareness | Letter Knowledge | Serial Recall | Oral Comprehension | Vocabulary | Grammar | Visual Short Term Memory | Nonverbal Ability | Word Decoding | Precursor Literacy |
|  | Phonem* Awareness | Grapheme Knowledge |  | Verbal Comprehension | Word  Knowledge | Syntax | Phonological Short Term Memory | Nonverbal Intelligence | Reading Fluency | Precursor Reading |
|  |  | Phoneme Grapheme Correspondence |  | Listening Comprehension |  | Syntac* | Short Term Memory | Nonverbal IQ | Word Recognition | Predictor Literacy |
|  |  | Sound Symbol* |  |  |  | Synta* | Working Memory |  | Reading Ability | Predictor Reading |
|  |  |  |  | |  | Morpholog* | Visual Memory |  | Reading Skills | Precursor Literacy Skills |
|  |  |  |  |  |  | Morphem* | Verbal Memory |  | Literacy Skills | Precursors of Reading Ability |
| **Literacy Precursors** | **Phonological Awareness** | **Letter Knowledge** | **RAN/**  **Serial Recall** | **Oral Language/ Listening Comprehension** | **Vocabulary** | **Grammar** | **Memory** | **Non-Verbal Intelligence** | **Word Decoding** | **General Literacy**  **Precursors** |
| **Search Query** |  |  |  | |  |  | Nonverbal Memory |  |  | Early Predictors of Later Conventional Literacy Skills |
|  |  |  |  |  |  |  |  |  |  | Predictive Literacy Skills Predictors of Later Reading Skills; Preschool Literacy |
| **Literacy Outcomes** | | | **Word/Non-Word Decoding Or Reading** | | | **Reading Comprehension** | | | |  |
| **Search Query** | | | Word Decoding  Reading Fluency | | | Reading Comprehension | | | | |
|  | | | Word Recognition | | |  |  |  |  |  |
|  |  |  | Reading Ability | | |  |  |  |  |  |
|  |  |  | Reading Skills | | |  |  |  |  |  |
|  |  |  | Literacy Skills | | |  |  |  |  |  |
| **Participant Demographic Characteristics** | | | **Age Demographic (Children)** | | | **Type of Language Background (Multilinguals/Bilinguals)** | | | | |
| **Search Query** | | | Child*  Infants | | | Bilingual*  Multilingual*  Second Language Learner* | | | | |

*Note. ** = truncation symbol to retrieve multiple search terms with a common root in the electronic databases.
