## Supplemental Table 4 for "Early Precursors of Literacy Development in Simultaneous Bilinguals: A Systematic Review and Meta-Analysis"

**S4**

*Studies with Assigned Study Numbers Included in the Systematic Literature Review and Meta-Analysis.* Table indicating the databases, assigned study numbers and statistical methods for the 41 study citations. The study numbers in Tables 1-5, and supplemental files S1-S18 will refer to the corresponding citations in S4.

| **Database** | **Reference** | **In-Text Citation** | | | | | **Statistics** | | | **Assigned Study Number** |
| --- | --- | --- | --- | --- | --- | --- | --- | --- | --- | --- |
| LLBA | Hipfner-Boucher, K., Lam, K., & Chen, X. (2014). The effects of bilingual education on the English language and literacy outcomes of Chinese-speaking children. *Written Language and Literacy, 17*(1), 116-138. https://doi.org/10.1075/wll.17.1.06hip | Hipfner-Boucher et al., 2014 | | | | | Pearson’s, hierarchical multiple regression (for some variables) | | | 1 |
| LLBA | Yeong, S. H. M., Fletcher, J., & Bayliss, D. M. (2014). Importance of phonological and orthographic skills for English reading and spelling: A comparison of English monolingual and Mandarin-English bilingual children. *Journal of Educational Psychology, 106*(4), 1107. https://psycnet.apa.org/doi/10.1037/a0036927 | Yeong et al., 2014 | | | | | Pearson’s, hierarchical multiple regression | | | 2 |
| LLBA | Hsu, L. S., Ip, K. I., Arredondo, M. M., Tardif, T., & Kovelman, I. (2016). Simultaneous acquisition of English and Chinese impacts children's reliance on vocabulary, morphological and phonological awareness for reading in English. *International Journal of Bilingual Education and Bilingualism, 22*(2), 207-223. http://dx.doi.org/10.1080/13670050.2016.1246515 | Hsu et al., 2016 | | | | | Pearson’s, path analysis | | | 3 |
| MLA; PsycINFO | Chiappe, P., Siegel, L. S., & Gottardo, A. (2002). Reading-related skills of kindergartners from diverse linguistic backgrounds. *Applied Psycholinguistics, 23*(1), 95-116. http://dx.doi.org/10.1017/S014271640200005X | Chiappe et al., 2002 | | | | | Pearson’s, stepwise multiple regression | | | 4 |
| MLA | Zhang, D. (2015). Morphology in Malay-English biliteracy acquisition: An intervention study. *International Journal of Bilingual Education and Bilingualism, 19*(5), 546-562. http://dx.doi.org/10.1080/13670050.2015.1026873 | Zhang, 2015 | | | | | ANOVA (effect size, Cohen’s *d*) | | | 5 |
| PsycINFO | Tamis-LeMonda, C., Song, L., Luo, R., Kuchirko, Y., Kahana-Kalman, R., Yoshikawa, H., & Raufman, J. (2014). Children's vocabulary growth in English and Spanish across early development and associations with school readiness skills. *Developmental Neuropsychology, 39*(2), 69-87. http://dx.doi.org/10.1080/87565641.2013.827198 | Tamis-LeMonda et al., 2014 | | | | | | | Multiple regression | 6 |
| PsycINFO | Jasińska, K.K., & Petitto, L. (2017). Age of bilingual exposure is related to the contribution of phonological and semantic knowledge to successful reading development. *Child Development, 89*(1), 310-331. http://dx.doi.org/10.1111/cdev.12745 | Jasińska & Petitto, 2017 | | | | | | | Multigroup structural equation modelling | 7 |
| PsycINFO | O'Brien, B. A., Mohamed, M. B. H., Yussof, N. T., & Ng, S. C. (2019). The phonological awareness relation to early reading in English for three groups of simultaneous bilingual children. *Reading and Writing: An Interdisciplinary Journal, 32*(4), 909-937. http://dx.doi.org/10.1007/s11145-018-9890-1 | O'Brien et al., 2019 | | | | | | | Pearson’s, multiple regression, linear regression | 8 |
| LLBA | Ibrahim, R., Eviatar, Z., & Aharon-Peretz, J. (2007). Metalinguistic awareness and reading performance: A cross language comparison. *Journal of Psycholinguistic Research, 36*(4), 297-317. https://link.springer.com/article/10.1007/s10936-006-9046-3 | Ibrahim et al., 2007 | | | | | | | Pearson’s, multiple regression | 9 |
| LLBA; PsycINFO | Hammer, C.S., Lawrence, F. R., & Miccio, A. W. (2007). Bilingual children's language abilities and early reading outcomes in head start and kindergarten. *Language, Speech & Hearing Services in Schools, 38*(3), 237-48. https://doi.org/10.1044/0161-1461(2007/025) | Hammer et al., 2007 | | | | | | | Growth curve modelling | 10 |
| LLBA; PsycINFO; ERIC | Dunn, D.M., Hammer, C., & Lawrence, F. R. (2011). Associations between preschool language and first grade reading outcomes in bilingual children. *Journal of Communication Disorders, 44*(4), 444-458. https://doi.org/10.1016/j.jcomdis.2011.02.003 | Dunn et al., 2011 | | | | | | | Growth curve modelling | 11 |
| LLBA | Oller, D. K., Pearson, B. Z., & Cobo-lewis, A. (2007). Profile effects in early bilingual language and literacy. *Applied Psycholinguistics, 28*(2), 191. http://dx.doi.org/10.1017/S0142716407070117 | Oller et al., 2007 | | | | | | | Scheffe test/MANOVA (effect size, Cohen’s *d*) | 12 |
| LLBA; ERIC | Bengochea, A., Justice, L. M., & Hijlkema, M. J. (2015). Print knowledge in Yucatec Maya-Spanish bilingual children: An initial inquiry. *International Journal of Bilingual Education and Bilingualism, 20*(7), 807-822. http://dx.doi.org/10.1080/13670050.2015.1103699 | | Bengochea et al., 2015 | | | | | Pearson’s | | 13 |
| PsycINFO | Cherodath, S., & Singh, N. C. (2015). The influence of orthographic depth on reading networks in simultaneous biliterate children. *Brain and Language, 143,* 42-51. http://dx.doi.org/10.1016/j.bandl.2015.02.001 | | Cherodath & Singh, 2015 | | | | | Repeated measures ANOVA | | 14 |
| PsycINFO | Lallier, M., Valdois, S., Lassus-Sangosse, D., Prado, C., & Kandel, S. (2014). Impact of orthographic transparency on typical and atypical reading development: Evidence in French-Spanish bilingual children. *Research in Developmental Disabilities, 35*(5), 1177-1190. http://dx.doi.org/10.1016/j.ridd.2014.01.021 | | Lallier et al., 2014 | | | | | Pearson’s | | 15 |
| Additional Search | Bérubé, D., & Marinova-Todd, S. (2011). The development of language and reading skills in the second and third languages of multilingual children in French immersion. *International Journal of Multilingualism 9*: 272-293. https://doi.org/10.1080/14790718.2011.631708 | | Bérubé & Marinova-Todd, 2011 | | | | | Pearson’s, hierarchical multiple regression | | 16 |
| Additional Search | D’angiulli, A., Siegel, L. S., & Serra. E. (2002). The development of reading in English and Italian in bilingual children. *Applied Psycholinguistics,* *22* (4): 479-507. https://doi.org/10.1017/S0142716401004015 | | D’angiulli et al., 2002 | | | | | Pearson’s | | 17 |
| Additional Search | Ellis, N. C., & Hooper, A. M. (2002). Why learning to read is easier in Welsh than in English: Orthographic transparency effects evinced with frequency-matched tests. *Applied Psycholinguistics, 22,* 571-599. https://doi.org/10.1017/S0142716401004052 | | Ellis & Hooper, 2002 | | | | | Multiple regression | | 18 |
| Additional Search | Jasińska, K. K., Wolf, S., Jukes, M. C., & Dubeck, M. M. (2019). Literacy acquisition in multilingual educational contexts: Evidence from Coastal Kenya. *Developmental Science, 128*(28). https://doi.org/10.1111/desc.12828 | | Jasińska et al., 2019 | | | | | Multiple regression | | 19 |
| Additional Search | Kovelman, I., Salah-Ud-Din, M., Berens, M. S., & Petitto, L. (2015). “One glove does not fit all” in bilingual reading acquisition: Using the age of first bilingual language exposure to understand optimal contexts for reading success. *Cogent Education, 2*(1). https://doi.org/10.1080/2331186X.2015.1006504 | | | Kovelman et al., 2015 | | Repeated measures MANOVA | | | | 20 |
| Additional Search | Lervåg, A., & Aukrust, V. G. (2010). Vocabulary knowledge is a critical determinant of the difference in reading comprehension growth between first and second language learners.” *Journal of Child Psychology and Psychiatry, 51* (5): 612-620.  https://doi.org/10.1111/j.1469-7610.2009.02185.x | | | Lervåg & Aukrust, 2010 | | Hierarchical multiple regression | | | | 21 |
| Additional Search | Spencer, L.H., & Hanley, J.R. (2010). Effects of orthographic transparency on reading and phoneme awareness in children learning to read in Wales. *British Journal of Psychology, 94:* 1-28. https://doi.org/10.1348/000712603762842075 | | | Spencer & Hanley, 2010 | | Pearson’s, stepwise multiple regression | | | | 22 |
| Additional Search | Lam, K., Chen, X., Geva, E., Luo, Y. C., & Li, H. (2012). The role of morphological awareness in reading achievement among young Chinese-speaking English language learners: A longitudinal study. *Reading and Writing*, *25*(8), 1847-1872. https://psycnet.apa.org/doi/10.1007/s11145-011-9329-4 | | | Lam et al., 2012 | | Pearson’s, hierarchical multiple regression | | | | 23 |
| Additional Search | Limbird, C. K., Maluch, J. T., Rjosk, C., Stanat, P., & Merkens, H. (2014). Differential growth patterns in emerging reading skills of Turkish-German bilingual and German monolingual primary school students. *Reading and Writing*, *27*(5), 945-968. https://psycnet.apa.org/doi/10.1007/s11145-013-9477-9 | | | Limbird et al., 2014 | | Pearson’s, multigroup structural equation modelling | | | | 24 |
| PsycINFO | Rhys, M., & Thomas, E. M. (2012). Bilingual Welsh-English children's acquisition of vocabulary and reading: Implications for bilingual education. *International Journal of Bilingual Education and Bilingualism, 16*(6), 633-656. http://dx.doi.org/10.1080/13670050.2012.706248 | | | Rhys & Thomas, 2012 | | Pearson’s | | | | 25 |
| LLBA | Spatgens, T., & Schoonen, R. (2017). The semantic network, lexical access, and reading comprehension in monolingual and bilingual children: An individual differences study. *Applied Psycholinguistics, 39*(1), 225-256. http://dx.doi.org/10.1017/S0142716417000224 | | | Spatgens & Schoonen, 2017 | | Fixed and random effects estimate analysis | | | | 26 |
| Additional Search | Gupta, A., & Jamal, G. (2007). Reading strategies of bilingual normally progressing and dyslexic readers in Hindi and English. *Applied Psycholinguistics, 28*(1), 47-68. https://doi.org/10.1017/S0142716406070032 | | | | Gupta & Jamal, 2007 | | ANOVA | | | 27 |
| Additional Search | Ríos-López, P., Molnar, M. T., Lizarazu, M., & Lallier, M. (2017). The role of slow speech amplitude envelope for speech processing and reading development. *Frontiers in Psychology*, *8*, 1497. https://doi.org/10.3389/fpsyg.2017.01497 | | | | Ríos-López et al., 2017 | | Pearson’s | | | 28 |
| LLBA | Vital, H., & Karniol, R. (2010). Procedural versus narrative cross-language priming and bilingual children's reading and sentence sequencing of same genre and opposite genre text in the other language. *Bilingualism, 14*(4), 547-561. http://dx.doi.org/10.1017/S1366728910000520 | | | | Vital & Karniol, 2010 | | ANOVA | | | 29 |
| LLBA; PsycINFO | van den Bosch, L. J., Segers, E., & Verhoeven, L. (2020). First and second language vocabulary affect early second language reading comprehension development. *Journal of Research in Reading, 43*(3), 290-308. https://doi.org/10.1111/1467-9817.12304 | | | | van den Bosch et al., 2020 | | Pearson’s, ANOVA | | | 30 |
| Additional Search: Research Callout | Sun, H., Bornstein, M.H., & Esposito, G. (2021). The specificity principle in young dual language learners’ English development. *Child Development.* <https://doi.org/10.1111/cdev.13558> | | | | Sun et al., 2021 | | LASSO | | | 31 |
| LLBA | Yang, F.Y. (2010). Biliteracy effects on phonological awareness, oral language proficiency and reading skills in Taiwanese Mandarin-English bilingual children. *ProQuest Dissertations & Theses Global* (276360261). https://citeseerx.ist.psu.edu/viewdoc/download?doi=10.1.1.1028.5605&rep=rep1&type=pdf | | | | Yang, 2010 | | Hierarchical multiple regression; ANOVA/MANOVA (effect size, Cohen’s *d*) | | | 32 |
| LLBA | Mak, K.C.L. (2013). Reading comprehension in Chinese-English bilingual children: A cognitive perspective (dissertation). *Library and Archives Canada = Bibliothèque et Archives Canada*. https://www.bac-lac.gc.ca/eng/services/theses/Pages/item.aspx?idNumber=921571256 | | | | Mak, 2013 | | Pearson’s, hierarchical multiple regression, Cohen’s *d* (effect size) | | | 33 |
| LLBA | Lallier, M., Martin, C. D., Acha, J., & Carreiras, M. (2021). Cross-linguistic transfer in bilingual reading is item specific. *Bilingualism: Language and Cognition*, *24*(5), 891-901. https://doi.org/10.1017/S1366728921000183 | | | | Lallier et al., 2021 | | Cohen’s *d* (effect size) for non-verbal intelligence only, ANCOVA, Pearson’s | | | 34 |
| LLBA | Sun, X., Zhang, K., Marks, R. A., Nickerson, N., Eggleston, R. L., Yu, C. L., Chou, T-L., Tardif,T., & Kovelman, I. (2022). What’s in a word? Cross‐linguistic influences on Spanish–English and Chinese–English bilingual children’s word reading development. *Child Development*, *93*(1), 84-100. https://srcd.onlinelibrary.wiley.com/doi/epdf/10.1111/cdev.13666 | | | | Sun et al., 2022 | | ANCOVA, multiple regression, partial correlations, multi-group path models | | | 35 |
| LLBA | Novita, S., Lockl, K., & Gnambs, T. (2022). Reading comprehension of monolingual and bilingual children in primary school: the role of linguistic abilities and phonological processing skills. *European Journal of Psychology of Education*, 1-21. https://doi.org/10.1007/s10212-021-00587-5 | | | | Novita et al., 2022 | | Cohen’s *d* (effect size), multiple regression | | | 36 |
| LLBA | Peets, K. F., Yim, O., & Bialystok, E. (2019). Language proficiency, reading comprehension and home literacy in bilingual children: The impact of context. *International Journal of Bilingual Education and Bilingualism*, *25*(1), 226-240. https://doi.org/10.1080/13670050.2019.1677551 | | | | Peets et al., 2019 | | ANOVA, multiple regression | | | 37 |
| LLBA | Ruan, Y., Ye, Y., Lui, K. F. H., McBride, C., & Ho, C. S. H. (2022). How Do Word Reading and Word Spelling Develop Over Time? A Three‐Year Longitudinal Study of Hong Kong Chinese–English Bilingual Children. *Reading Research Quarterly*. https://doi.org/10.1002/rrq.478 | | | | Ruan et al., 2022 | | Pearson’s, cross-lagged panel model analysis | | | 38 |
| LLBA; MLA | Babayiğit, S., Hitch, G. J., Kandru-Pothineni, S., Clarke, A., & Warmington, M. (2022). Vocabulary limitations undermine bilingual children’s reading comprehension despite bilingual cognitive strengths. *Reading and Writing*, 1-23.  https://doi-org.myaccess.library.utoronto.ca/10.1007/s11145-021-10240-8 | | | | Babayiğit et al., 2022 | | Pearson’s, ANCOVA, path analysis | | | 39 |
| LLBA | Marks, R. A., Sun, X., McAlister López, E., Nickerson, N., Hernandez, I., Caruso, V. C., Satterfield, T., & Kovelman, I. (2022). Cross-linguistic differences in the associations between morphological awareness and reading in Spanish and English in young simultaneous bilinguals. *International Journal of Bilingual Education and Bilingualism*, *25*(10), 3907-3923. <https://doi.org/10.1080/13670050.2022.2090226> | | | | Marks et al., 2022 | | Pearson’s, Path analysis | | | 40 |
| PsycINFO | Gunnerud, H. L., Foldnes, N., & Melby-Lervåg, M. (2022). Levels of skills and predictive patterns of reading comprehension in bilingual children with an early age of acquisition. *Reading and Writing*, 1-23. https://doi.org/10.1007/s11145-022-10286-2 | | | | Gunnerud et al., 2022 | | Pearson’s, Multiple regression; Multigroup confirmatory factor analysis | | | 41 |

*Note.* LLBA= Linguistics and Language Behavior Abstracts database; ERIC= Educational Resources Informational Center database, MLA= MLA International Bibliography database; Additional Search = Google Scholar and manual citation search.
