## Supplemental Table 5 for "Early Precursors of Literacy Development in Simultaneous Bilinguals: A Systematic Review and Meta-Analysis"

**S5**

*Extracted Data Items Corresponding to Studies Included in the Systematic Literature Review and Meta-Analysis.* Summary table indicating the bilingual language groups, type of literacy precursor, testing medium, languages and measures of literacy precursor assessment, type of literacy outcome, languages and measures of literacy outcome assessment, mean age at assessment, sample size, and country of assessment corresponding to the 41 studies listed in S4.

| **Study** | **Bilingual Language Group** | **Literacy Precursors** | **Testing Medium** | **Languages and Measures of Literacy Precursor Assessment** | **Literacy Outcomes** | **Languages and Measures of Literacy Outcome Assessment** | **Mean Age**  **(in years)** | **n** | **Country of Assessment** |
| --- | --- | --- | --- | --- | --- | --- | --- | --- | --- |
| **1** | Chinese-English | Phonological Awareness | Within; Across | Chinese (phoneme and syllable deletion); English (Comprehensive Test of Phonological Processing; CTOPP; phoneme and syllable deletion) | Word Reading Accuracy | English (Letter-Word Identification subtest, Woodcock Language Proficiency Battery; WLPB) |  |  | Canada |
|  |  | Receptive Vocabulary | Within | English (Peabody Picture Vocabulary Test; PPVT) | Word Reading Accuracy | English (Letter-Word Identification subtest; WLPB) |  |  |  |
|  |  | Morphological Awareness | Within | English (Compound word awareness) | Word Reading Accuracy | English (Letter-Word Identification subtest; WLPB) |  |  |  |
| *1A* | Bilingual Chinese-English schooling |  |  |  |  |  | 6.10 | 20 |  |
| *1B* | English-only schooling |  |  |  |  |  | 6.07 | 33 |  |
| **2** | Chinese-English | Phonological Awareness | Within | English (CTOPP: Silent Phonological Choice Task); phoneme and syllable blending; phonological recording | Word Reading Accuracy | English (Wechsler Individual Achievement Test 2: Australian Adaptation; WIAT) |  |  | Singapore |
|  |  | Orthographic Processing Skills | Within | English (orthographic choice task; homophone verification task; non-lexical choice task) | Word Reading Accuracy | English (WIAT) |  |  |  |
| *2A* | Grade 2-3 English L1 |  |  |  |  |  | 8.02 | 29 |  |
| *2B* | Grade 2-3 Mandarin L1 |  |  |  |  |  | 8.02 | 27 |  |
| *2C* | Grade 5-6 English L1 |  |  |  |  |  | 11 | 25 |  |
| *2D* | Grade 5-6 Mandarin L1 |  |  |  |  |  | 11 | 25 |  |
| **3** | Chinese-English | Phonological Awareness | Within; Across | Chinese (phoneme and syllable deletion); English (CTOPP; phoneme and syllable deletion) | Word Reading Accuracy | Chinese (Character recognition test); English (Word Identification subtest of the Woodcock Reading Mastery Tests – Revised; WRMT-R) | 8.07 | 57 | US |
|  |  | Receptive Vocabulary | Within; Across | Chinese (Cantonese Receptive Vocabulary Test; CRVT); English (vocabulary subtest, Kaufman Brief Intelligence Test, Second Edition; KBIT-2) | Word Reading Accuracy | Chinese (Character recognition test); English (Word Identification subtest; WRMT-R) |  |  |  |
|  |  | Morphological Awareness | Within; Across | Chinese (Morphological Construction Judgement Task); English (Test of Morphological Structure) | Word Reading Accuracy | Chinese (Character recognition test); English (Word Identification subtest; WRMT-R) |  |  |  |
| **4** | Heterogenous (Farsi, Japanese, Spanish, Tagalog, Chinese, French, Slovakian, Squamish, Arabic, German, Greek, Hindi or Indonesian) - English | Phonological Awareness | Within | English (sound mimicry, rhyme detection, syllable and phoneme identification, and phoneme deletion) | Word Reading Accuracy | English (letter-word reading subtest of the Wide Range Achievement Test–3 ; WRAT-3) | 5.04 | 59 | Canada |
|  |  | Verbal Short-Term Memory | Within | English (Memory for Sentences subtest of the Stanford Binet Test) | Word Reading | English (letter-word reading subtest; WRAT-3) |  |  |  |
|  |  | Syntactic Awareness | Within | English (oral cloze task) | Word Reading Accuracy | English (letter-word reading subtest; WRAT-3) |  |  |  |
|  |  | Letter Knowledge | Within | English (letter identification task) | Word Reading Accuracy | English (letter-word reading subtest; WRAT-3) |  |  |  |
|  |  | RAN/Serial Recall | Within | English (rapid picture naming) | Word Reading Accuracy | English (letter-word reading subtest; WRAT-3) |  |  |  |
|  |  | Spelling | Within | English (word spelling) | Word Reading Accuracy | English (letter-word reading subtest; WRAT-3) |  |  |  |
|  |  | Environmental Print Awareness | Within | English (naming of common English logos) | Word Reading Accuracy | English (letter-word reading subtest; WRAT-3) |  |  |  |
| **5** | Malay-English | Morphological Awareness | Within; Across | English | Word Reading Accuracy; Word Reading Fluency | English; Malay | 9.04 | 64 | Singapore |
| **6** | Spanish-English | Expressive Vocabulary | Within; Across | English (vocabulary growth (word type/minute) during book-sharing sessions); Spanish (vocabulary growth (word type/minute) during book-sharing sessions) | Word Reading Accuracy | English (letter-word identification subtest, Woodcock Johnson III Tests of Achievement; WJ-III) | 2-5 | 133 | US |
| **7** | Heterogenous (Spanish, Tamil, Arabic, Hungarian, Urdu, or Chinese)- English ; French-English | Phonological Awareness* | Within | English (initial phoneme deletion, final phoneme deletion and phoneme segmentation) | Word Reading Accuracy | English (WLPB-R) | 8.05 | 139 | Canada |
|  |  | Semantic Awareness | Within | English (passage comprehension, synonym generation and antonym generation; Woodcock Language Proficiency Battery–Revised | Word Reading Accuracy | English (WLPB-R) |  |  |  |
| **8** | Malay-English; Tamil-English; Mandarin- English | Phonological Awareness | Within | English (CTOPP: onset-rime, syllable and phoneme deletion and blending) | Word Reading Accuracy | English (Wide Range Achievement Test; WRAT-4) |  |  | Singapore |
|  |  | Receptive Vocabulary | Across | Mandarin(8A)/Malay(8B)/Tamil(8C): Bilingual Language Assessment Battery (BLAB) | Word Reading Accuracy | English (WRAT-4) |  |  |  |
| *8A* | Mandarin-English |  |  |  |  |  | 4.08-5.08 | 311 |  |
| *8B* | Malay-English |  |  |  |  |  | 4.08-5.08 | 147 |  |
| *8C* | Tamil-English |  |  |  |  |  | 4.08-5.08 | 163 |  |
| **9** | Russian-Hebrew | Phonological Awareness | Within | Hebrew (initial phoneme detection, final phoneme detection, and syllable/phoneme detection) | Word Reading Errors; Non-Word Reading Accuracy Errors; Text Reading Errors; Text Reading Fluency | Hebrew | 6.10-7.03 | 19 | Middle-East |
|  |  | Semantic Awareness | Within | Hebrew (WISC-R: word definition test) | Word Reading Errors; Non-Word Reading Accuracy Errors; Text Reading Errors; Text Reading Fluency | Hebrew |  |  |  |
| **10** | Spanish-English | Receptive Vocabulary and Oral Language Comprehension | Within; Across | Spanish(Test de vocabulario en ima´genes Peabody; TVIP and Preschool Language Scale-3; PLS-3); English (PPVT-III and Test of Early Language Development- 3; TELD-3) | Word Reading Accuracy | Spanish (WLPB-R); English (WLPB-R) | 3.09 | 53 | US |
| **11** | Spanish-English | Receptive Vocabulary | Within; Across | Spanish (TVIP); English (PPVT-III) | Word Reading Accuracy; Text Reading Comprehension | Spanish (letter–word identification and passage comprehension subtests; WLPB-R); English (letter–word identification and passage comprehension subtests; WLPB-R) | 3.08 | 48 | US |
|  |  | Oral Language Comprehension | Within; Across | Spanish (receptive language subtest; PLS-3); English (auditory comprehension subtest; TELD-3) | Word Reading Accuracy; Text Reading Comprehension | Spanish (letter–word identification and passage comprehension subtests; WLPB-R); English (letter–word identification and passage comprehension subtests; WLPB-R) |  |  |  |
| **12** | Spanish-English | Type of Literacy Outcome Measure | Within | Spanish (Woodcock–Johnson and Woodcock–Munoz language and literacy evaluations); English (Woodcock–Johnson and Woodcock–Munoz language and literacy evaluations) | Non-Word Reading Accuracy | Spanish (Woodcock–Johnson and Woodcock–Munoz language and literacy evaluations); English (Woodcock–Johnson and Woodcock–Munoz language and literacy evaluations) |  |  | US |
| *12A* | Grade 2 |  |  |  |  |  | Grade 2 | 704 |  |
| *12B* | Grade 5 |  |  |  |  |  | Grade 5 | 704 |  |
| **13** | Maya-Spanish | Letter Knowledge | Within; Across | Spanish (Phonological Awareness Literacy Screening-PreK; PALS); Maya | Word Reading Accuracy | Spanish; Maya | 4.10 | 84 | Spain |
|  |  | Name Writing | Across | Spanish; Maya | Word Reading Accuracy | Spanish; Maya |  |  |  |
| **14** | Hindi-English | Phonological Awareness (Neural Study) | Within; Across | Hindi; English | Word Reading Accuracy; Non-Word Reading Accuracy | Hindi; English | 9.21 | 44 | India |
| **15** | Spanish-French | Phonological Awareness* | Within | Spanish (initial phoneme deletion and acronym task); French (initial phoneme deletion and acronym task) | Non-Word Reading Accuracy; Text Reading Fluency | Spanish; French | 10.04 | 9 | Spain |
|  |  | Visual Attention (VA) Span | Within | Spanish and French (whole report task and partial report task) | Word Reading Accuracy; Word Reading Fluency; Non-Word Reading Accuracy; Non-Word Reading Fluency; Text Reading Errors; Text Reading Fluency | Spanish; French |  |  |  |
| **16** | Alphabetic (Africans, Amharic, Croatian, Czech, Danish, Fanti, German, Greek, Hungarian, Korean, Polish, Serbian, Spanish, Tagalog or Vietnamese ) - English;  Logographic (Cantonese, Japanese, Mandarin Chinese, Shanghainese ) -English | Receptive Vocabulary | Within | English (PPVT-III) | Word Reading Accuracy; Non-Word Reading Accuracy; Text Reading Comprehension | English (Woodcock Language Battery Proficiency; WLPB-R) |  |  | Canada |
|  |  | Oral Language Comprehension | Within | English (Listening Comprehension subtest, Woodcock Language Battery Proficiency; WLPB-R) | Word Reading Accuracy; Non-Word Reading Accuracy; Text Reading Comprehension | English (Woodcock Language Battery Proficiency; WLPB-R) |  |  |  |
| *16A* | Alphabetic L1 |  |  |  |  |  | 9 | 26 |  |
| *16B* | Logographic L1 |  |  |  |  |  | 9 | 13 |  |
| **17** | Italian-English | Syntactic Awareness | Within; Across | Italian (oral cloze); English (oral cloze) | Word Reading Accuracy; Non-Word Reading Accuracy | Italian; English (reading subtest; WRAT-R and Word Attack subtest; WRMT-R) | 9-13 | 81 | Canada |
|  |  | Working Memory | Within; Across | Italian (missing word repetition); English (missing word repetition) | Word Reading Accuracy; Non-Word Reading Accuracy | Italian; English (reading subtest; WRAT-R and Word Attack subtest; WRMT-R) |  |  |  |
|  |  | Spelling | Within; Across | Italian; English (Spelling sub-test, Wide Range Achievement Test–Revised; WRAT-R) | Word Reading Accuracy; Non-Word Reading Accuracy | Italian; English (reading subtest; WRAT-R and Word Attack subtest; WRMT-R) |  |  |  |
| **18** | Welsh-English | Type of Literacy Outcome Measure | Across | Welsh; English | Word Reading Accuracy; Word Reading Errors; Word Reading Fluency; Text Reading Comprehension | Welsh; English | 6-7 | 20 | Wales |
| **19** | Heterogenous-Kiswahili | Phonological Awareness* | Within; Across | Kiswahili (initial sound matching) | Word Reading Fluency | Kiswahili; English | 6.8-8.6 | 977 | Kenya |
|  |  | Receptive Vocabulary | Within; Across | Kiswahili (word picture matching) | Word Reading Fluency | Kiswahili; English |  |  |  |
| **20** | Spanish-English | Type of Reading Instruction | Within | Spanish; English (Phonics Reading Instruction; Success for All; SFA and Whole Language Reading Instruction) | Word Reading Accuracy; Non-Word Reading Accuracy; Text Reading Comprehension | Spanish; English | 7-9 | 23 | US |
| **21** | Urdu-Norwegian | Receptive Vocabulary | Within | Norwegian (PPVT-III and Wechsler Intelligence Scale for Children III; WISC III) | Text Reading Comprehension | Norwegian (passage comprehension subtest; WRMT-PC and the Neale Analysis of Reading Ability II ; NARA II) | 7.6 | 90 | Norway |
|  |  | Word Decoding | Within | Norwegian (Test of Word Reading Efficiency; TOWRE) | Text Reading Comprehension | Norwegian (passage comprehension subtest; WRMT-PC and NARA II) |  |  |  |
| **22** | Welsh-English | Phonological Awareness* | Within; Across | Welsh (phoneme segmentation); English (phoneme segmentation) | Word Reading Accuracy | Welsh; English | 6.01-7.0 | 74 | Wales |
|  |  | Receptive Vocabulary | Within; Across | Welsh (British Picture Vocabulary Scale; BPVS) | Word Reading Accuracy | Welsh; English |  |  |  |
|  |  | RAN/Serial Recall | Within; Across | Welsh and English (Digit span test, WISC-R) | Word Reading Accuracy | Welsh; English |  |  |  |
|  |  | Non-Verbal Intelligence | Within; Across | Welsh and English (Raven’s Coloured Progressive Matrices; CPM) | Word Reading Accuracy | Welsh; English |  |  |  |
| **23** | Chinese-English | Phonological Awareness* | Within | English (CTOPP-2: phoneme deletion) | Word Reading Accuracy; Text Reading Comprehension | English (Reading Comprehension subtest, Peabody Individual Achievement Test— Revised; PIAT-R) |  |  | Canada |
|  |  | Receptive Vocabulary | Within | English (PPVT-III) | Word Reading Accuracy; Text Reading Comprehension | English (Reading Comprehension subtest; PIAT-R) |  |  |  |
|  |  | Morphological Awareness | Within | English (Derivational Awareness task and Compound Awareness Task | Word Reading Accuracy; Text Reading Comprehension | English (Reading Comprehension subtest; PIAT-R) |  |  |  |
|  |  | Word Decoding | Within | English (Letter-Word Identification subtest; WLPB) | Text Reading Comprehension | English (Reading Comprehension subtest; PIAT-R) |  |  |  |
| *23A* | Kindergarten |  |  |  |  |  | 5.05 | 46 |  |
| *23B* | Grade 1 |  |  |  |  |  | 6.07 | 34 |  |
| **24** | Turkish-German | Phonological Awareness* | Within | German (Basiskompetenzen fu¨r Lese-Rechtschreibleistungen; BAKO 1–4: phoneme identification, deletion, word remainder determination and sound categorization) | Text Reading Comprehension | German Text Comprehension subtest, Ein Leseversta¨ndnistest fu¨r Elementarschu¨ler’; ELFE) | 7.09 | 100 | Germany |
|  |  | Expressive Vocabulary | Within | German (Bilingual Verbal Abilities Test; BVAT) | Text Reading Comprehension | German Text Comprehension subtest, Ein Leseversta¨ndnistest fu¨r Elementarschu¨ler’; ELFE) |  |  |  |
|  |  | Word Decoding | Within | German (Wu¨rzburg Silent Reading Test; WLLP) | Text Reading Comprehension | German Text Comprehension subtest, Ein Leseversta¨ndnistest fu¨r Elementarschu¨ler’; ELFE) |  |  |  |
| **25** | Welsh-English | Receptive Vocabulary | Within | Welsh Prawf Geirfa Cymraeg; PGC); English (BPVS) | Word Reading Accuracy; Text Reading Comprehension | Welsh (Profion Darllen Glannau Menai); English (NARA-II) | 7-11 | 38 | Wales |
| **26** | Heterogenous-Dutch | Receptive Vocabulary | Within | Dutch (Cito Leeswoordenschat) | Text Reading Comprehension | Dutch (Begrijpend Lezen 678) | 11.03 | 86 | Netherlands |
|  |  | Semantic Awareness | Within | Dutch (auditory semantic decision task) | Text Reading Comprehension | Dutch (Begrijpend Lezen 678) |  |  |  |
|  |  | Word Decoding | Within | Dutch (Drie Minuten Toets) | Text Reading Comprehension | Dutch (Begrijpend Lezen 678) |  |  |  |
|  |  | RAN/Serial Recall | Within | Dutch (Rapid Automatized Naming Test; RAN and Rapid Alternating Stimulus Test). | Text Reading Comprehension | Dutch (Begrijpend Lezen 678) |  |  |  |
| **27** | Hindi-English | Type of Literacy Outcome Measure | Within | Hindi; English | Word Reading Accuracy | Hindi; English | 8.06 | 30 | India |
| **28** | Basque-Spanish | Speech Perception | Within | Spanish | Word Reading Accuracy; Word Reading Errors; Non-Word Reading Accuracy; Non-Word Reading Errors; Text Reading Errors; Text Reading Fluency | Spanish (El principito and PROLEC-R) |  | 20 | Spain |
| *28A* | Grade 2 |  |  |  |  |  | 7.07 |  |  |
| *28B* | Grade 5 |  |  |  |  |  | 11 |  |  |
| **29** | Hebrew-English | Sentence Priming | Across | English | Text Reading Errors; Text Reading Fluency; Text Reading Comprehension | Hebrew | Grades 5-6 | 101 | Middle-East |
| **30** | Turkish-Dutch | Receptive Vocabulary | Within; Across | Turkish (PPVT); Dutch (T-TOS) | Word Reading Accuracy; Non-Word Reading Accuracy; Text Reading Comprehension | Dutch | 7.05 | 71 | Netherlands |
|  |  | Word Decoding | Within | Dutch | Text Reading Comprehension | Dutch |  |  |  |
|  |  | Non-Word Decoding | Within | Dutch | Text Reading Comprehension | Dutch |  |  |  |
| **31** | Chinese-English | Working Memory | Within | English (Backward Digit Recall) | Word Reading Accuracy | English (Word Reading Subtest; WRAT) | 4.01-5.05 | 736 | Singapore |
|  |  | Non-Verbal Intelligence | Within | English (Raven’s CPM) | Word Reading Accuracy | English (Word Reading Subtest; WRAT) |  |  |  |
| **32** | Chinese-English | Phonological Awareness | Within; Across | Chinese (onset-rime matching task and tone matching task); English (onset-rime matching task and final phoneme matching task) | Word Reading Accuracy | Chinese (Graded Character Recognition Task); English (Word Identification Subtest; WRAT-R) | 8.05 | 40 | US |
|  |  | Non-Verbal Intelligence | Within | English (Test of Nonverbal Intelligence, Third Edition; TONI-3) | Word Reading Accuracy | English (Word Identification Subtest; WRAT-R) |  |  |  |
| **33** | Chinese-English | Phonological Awareness | Within; Across | Chinese (onset-rime and syllable deletion); English (CTOPP: phoneme deletion) | Word Reading Accuracy; Non-Word Reading Accuracy; Text Reading Comprehension | Chinese (Character Recognition Test, Pseudo-Character Test and Gray Oral Reading Test Fourth Edition; GORT-4); English (Word Identification Subtest, WRMT-R, Word Attack Subtest; WRMT-R and NARA) | 11 | 47 | Canada |
|  |  | RAN/Serial Recall | Within; Across | Chinese (Forward Digit Span subtest, Wechsler Intelligence Scale for Children - Third Edition; WISC-III) and English (Forward Digit Span subtest; WISC-III) | Word Reading Accuracy; Non-Word Reading Accuracy; Text Reading Comprehension | Chinese (Character Recognition Test, Pseudo-Character Test and Gray Oral Reading Test Fourth Edition; GORT-4); English (Word Identification Subtest, WRMT-R, Word Attack Subtest; WRMT-R and NARA) |  |  |  |
|  |  | Receptive Vocabulary | Within; Across | Chinese (PPVT-III); English (PPVT-III) | Word Reading Accuracy; Non-Word Reading Accuracy; Text Reading Comprehension | Chinese (Character Recognition Test, Pseudo-Character Test and Gray Oral Reading Test Fourth Edition; GORT-4); English (Word Identification Subtest, WRMT-R, Word Attack Subtest; WRMT-R and NARA) |  |  |  |
|  |  | Expressive Vocabulary | Within; Across | Chinese (Expressive One-Word Picture Vocabulary Test - Third edition; EOWPVT-III); English  ( EOWPVT-III) | Word Reading Accuracy; Non-Word Reading Accuracy; Text Reading Comprehension | Chinese (Character Recognition Test, Pseudo-Character Test and Gray Oral Reading Test Fourth Edition; GORT-4); English (Word Identification Subtest, WRMT-R, Word Attack Subtest; WRMT-R and NARA) |  |  |  |
|  |  | Non-Verbal Intelligence | Within; Across | Chinese and English (Matrix Analogies Test) | Word Reading Accuracy; Non-Word Reading Accuracy; Text Reading Comprehension | Chinese (Character Recognition Test, Pseudo-Character Test and Gray Oral Reading Test Fourth Edition; GORT-4); English (Word Identification Subtest, WRMT-R, Word Attack Subtest; WRMT-R and NARA) |  |  |  |
|  |  | Word Decoding | Within; Across | Chinese (Chinese Character Recognition test); English (word identification subtest; WRMT-R) | Word Reading Accuracy; Non-Word Reading Accuracy; Text Reading Comprehension | Chinese (Character Recognition Test, Pseudo-Character Test and Gray Oral Reading Test Fourth Edition; GORT-4); English (Word Identification Subtest, WRMT-R, Word Attack Subtest; WRMT-R and NARA) |  |  |  |
|  |  | Non-Word Decoding | Within; Across | Chinese (Chinese Pseudo-Character Test); English (word attack subtest; WRMT-R) | Word Reading Accuracy; Non-Word Reading Accuracy; Text Reading Comprehension | Chinese (Character Recognition Test, Pseudo-Character Test and Gray Oral Reading Test Fourth Edition; GORT-4); English (Word Identification Subtest, WRMT-R, Word Attack Subtest; WRMT-R and NARA) |  |  |  |
|  |  | Verbal Working Memory/Serial Recall | Within; Across | Chinese (Backward Digit Span subtest; WISC-III and Verbal Working Memory Task); English (Backward Digit Span subtest; WISC-III and Verbal Working Memory Task) | Word Reading Accuracy; Non-Word Reading Accuracy; Text Reading Comprehension | Chinese (Character Recognition Test, Pseudo-Character Test and Gray Oral Reading Test Fourth Edition; GORT-4); English (Word Identification Subtest, WRMT-R, Word Attack Subtest; WRMT-R and NARA) |  |  |  |
|  |  | Non-Verbal Working Memory | Within; Across | Chinese and English (Swanson’s Visual Matrix subtest, Test of Working Memory) | Word Reading Accuracy; Non-Word Reading Accuracy; Text Reading Comprehension | Chinese (Character Recognition Test, Pseudo-Character Test and Gray Oral Reading Test Fourth Edition; GORT-4); English (Word Identification Subtest, WRMT-R, Word Attack Subtest; WRMT-R and NARA) |  |  |  |
|  |  | Syntactic Awareness | Within; Across | Chinese (Chinese Syntactic Awareness Task); English (Syntactic Judgment task) | Word Reading Accuracy; Non-Word Reading Accuracy; Text Reading Comprehension | Chinese (Character Recognition Test, Pseudo-Character Test and Gray Oral Reading Test Fourth Edition; GORT-4); English (Word Identification Subtest, WRMT-R, Word Attack Subtest; WRMT-R and NARA) |  |  |  |
|  |  | Morphological Awareness | Within; Across | Chinese (Chinese Derivational and Compound Morpheme Awareness Test); English (Test of Morphological Structure) | Word Reading Accuracy; Non-Word Reading Accuracy; Text Reading Comprehension | Chinese (Character Recognition Test, Pseudo-Character Test and Gray Oral Reading Test Fourth Edition; GORT-4); English (Word Identification Subtest, WRMT-R, Word Attack Subtest; WRMT-R and NARA) |  |  |  |
| **34** | Spanish-Basque; French-Basque | VA Span | Across | Spanish, French, and Basque (Visual 1 Back Task) | Word Reading Accuracy; Non-Word Reading Accuracy; Word Reading Fluency; Non-Word Reading Fluency | Basque simple (no consonant clusters) and complex (one or multiple consonant clusters or digraphs) word and non-words | Grade 3 |  | Spain |
| *34A* | Spanish-Basque |  |  |  |  |  | 8.88 | 25 |  |
| *34B* | French-Basque |  |  |  |  |  | 8.96 | 25 |  |
| **35** | Spanish-English; Chinese-English | Phonological Awareness | Within; Across | Chinese (Newman et al.’s 2011 measure; phoneme and syllable deletion); English (Comprehensive Test of Phonological Processing; CTOPP; phoneme and syllable deletion); Spanish (Test of  Phonological Processing in Spanish; phoneme and syllable deletion) | Word Reading Accuracy | English (letter-word identification subtest, Woodcock Johnson IV Tests of Achievement; WJ-IV) | Kindergarten-Grade 4 |  | US |
|  |  | Morphological Awareness | Within; Across | Chinese (adapted Song et al.’s 2015 measure); English (Early Lexical Morphology Measure); Spanish (modelled after English Early Lexical Morphology Measure) | Word Reading Accuracy | English (letter-word identification subtest, Woodcock Johnson IV Tests of Achievement; WJ-IV) |  |  |  |
|  |  | Receptive Vocabulary | Within; Across | Chinese (PPVT-R); English (PPVT-5); Spanish (PPVT) | Word Reading Accuracy | English (letter-word identification subtest, Woodcock Johnson IV Tests of Achievement; WJ-IV) |  |  |  |
| *35A* | Spanish-English |  |  |  |  |  |  | 96 |  |
| *35B* | Chinese-English |  |  |  |  |  |  | 86 |  |
| **36** | Heterogeneous-German | Phonological Awareness | Within | German (Onset-Rime test from Build the Right Word) | Text Reading Comprehension | German | Kindergarten-Grade 4 |  | Germany |
|  |  | Receptive Vocabulary | Within | German (modified PPVT) | Text Reading Comprehension | German |  |  |  |
|  |  | Receptive Grammar (Morphological awareness and syntactic awareness) | Within | German (adapted Test of Reception of Grammar; morphological awareness and syntactic awareness) | Text Reading Comprehension | German |  |  |  |
|  |  | RAN/Serial Recall | Within | German (digit recall subtest of the Kaufman Assessment Battery for Children) | Text Reading Comprehension | German |  |  |  |
|  |  | Working Memory | Within | German (digit span backward subtest from the Hamburg Wechsler Intelligence Test for Children-III) | Text Reading Comprehension | German |  |  |  |
| *36A* | German  (dominant)-Heterogenous |  |  |  |  |  |  | 130 |  |
| *36B* | Heterogeneous (dominant)-German |  |  |  |  |  |  | 90 |  |
| **37** | Heterogeneous (Mandarin, Korean,  Russian, Tamil, Farsi, Hebrew, Vietnamese, Polish, Gujarati, Spanish, Portuguese, Serbian, and  Tagalog)-English | Nonverbal Verbal Intelligence | Within | English  (Matrices subtest of the Kaufmann’s Brief Intelligence Test, 2nd edition; K-BIT 2; visuo-spatial reasoning) | Word Reading Accuracy; Text Reading Comprehension | English (Word Identification subtest of the Woodcock Reading Mastery Tests – Revised - WRMT-R; Gates-MacGinitie Reading Test – Form C) | Grade 3  7.8-9.3 | 50 | Canada |
|  |  | Receptive Vocabulary | Within | English (PPVT-III) | Word Reading Accuracy; Text Reading Comprehension | English (Word Identification subtest of the Woodcock Reading Mastery Tests – Revised - WRMT-R; Gates-MacGinitie Reading Test – Form C) |  |  |  |
|  |  | Receptive Grammar  (Morphological awareness and syntactic awareness) | Within | English (Morphological and Syntactic awareness; Grammatical Completion subtest of the Test of Language Development: Primary; TOLD; morphological awareness and syntactic awareness) | Word Reading Accuracy; Text Reading Comprehension | English (Word Identification subtest of the Woodcock Reading Mastery Tests – Revised - WRMT-R; Gates-MacGinitie Reading Test – Form C) |  |  |  |
|  |  | Word Decoding | Within | English (Word Identification subtest of the Woodcock Reading Mastery Tests) | Text Reading Comprehension | English (Gates-MacGinitie Reading Test – Form C) |  |  |  |
| **38** | Chinese-English | Phonological Awareness (syllable deletion and onset deletion) | Within; Across (Chinese-English) | Chinese Phonological Awareness | Word Reading Accuracy | Chinese Word Decoding | Grades 1-3  7.38 | 364 | China |
|  |  | Orthographic Processing | Within | Chinese Orthographic Awareness; English (Word Decision Task) | Word Reading Accuracy | Chinese Word Reading Accuracy; English Word Reading Accuracy |  |  |  |
|  |  | Morphological Awareness | Within | Chinese Morphological Awareness; English Morphological Awareness | Word Reading Accuracy | Chinese Word Reading Accuracy; English Word Reading Accuracy |  |  |  |
|  |  | Expressive Vocabulary | Within | Chinese Expressive Vocabulary; English Expressive Vocabulary | Word Reading Accuracy | Chinese Word Reading Accuracy; English Word Reading Accuracy |  |  |  |
|  |  | Receptive Vocabulary | Within | Chinese Receptive Vocabulary; English Receptive Vocabulary | Word Reading Accuracy | Chinese Word Reading Accuracy; English Word Reading Accuracy |  |  |  |
|  |  | Spelling | Within | Chinese (Word Dictation Task); English (Word Dictation Task) | Word Reading Accuracy | Chinese Word Reading Accuracy; English Word Reading Accuracy |  |  |  |
|  |  | Nonverbal Verbal Intelligence | Within | Chinese and English (Raven’s CPM) | Word Reading Accuracy | Chinese Word Reading Accuracy; English Word Reading Accuracy |  |  |  |
|  |  | RAN/Serial Recall | Within | Chinese RAN; English RAN | Word Reading Accuracy | Chinese Word Reading Accuracy; English Word Reading Accuracy |  |  |  |
| **39** | Urdu-English | Expressive Vocabulary | Within | English (Wechsler Abbreviated Scale of Intelligence –II; WASI-II) | Word Reading Accuracy; Text Reading Comprehension | English (Single Word Reading Test; York Assessment of Reading for Comprehension) | Grades 4-6  9.06 | 104 | UK |
|  |  | Receptive Vocabulary | Within | English (British Picture Vocabulary Scale-3; BPVS-3) | Word Reading Accuracy; Text Reading Comprehension | English (Single Word Reading Test; York Assessment of Reading for Comprehension) |  |  |  |
|  |  | Nonverbal Verbal Intelligence | Within | English (Matrix Reasoning subtest of the WASI-II) | Word Reading Accuracy; Text Reading Comprehension | English (Single Word Reading Test; York Assessment of Reading for Comprehension) |  |  |  |
|  |  | Working Memory | Within | English (Backward Digit Span subtest of the Automated Working Memory Assessment) | Word Reading Accuracy; Text Reading Comprehension | English (Single Word Reading Test; York Assessment of Reading for Comprehension) |  |  |  |
|  |  | Executive Function | Within | English (Simon Task; Letter Fluency Task) | Word Reading Accuracy; Text Reading Comprehension | English (Single Word Reading Test; York Assessment of Reading for Comprehension) |  |  |  |
|  |  | Novel Word Learning | Within | English Novel Word Learning | Word Reading Accuracy; Text Reading Comprehension | English (Single Word Reading Test; York Assessment of Reading for Comprehension) |  |  |  |
|  |  | Word Decoding | Within | English (Single Word Reading Test) | Word Reading Accuracy; Text Reading Comprehension | English (York Assessment of Reading for Comprehension) |  |  |  |
| **40** | Spanish-English | Phonological Awareness) | Within; Across (Spanish-English) | Spanish (Test of Phonological Processing in Spanish; TOPPS; phoneme and syllable deletion); English (Comprehensive Test of Phonological Processing; CTOPP-2; phoneme and syllable deletion) | Word Reading Accuracy; Text Reading Comprehension | Spanish (Letter Word Identification and Passage Comprehension subtests of the Batería III Woodcock-Muñoz Test of Achievement); English (Letter Word Identification and Passage Comprehension subtests of the Woodcock-Johnson IV Test of Achievement) | 8.07 | 90 | US |
|  |  | Receptive Vocabulary | Within; Across (Spanish-English) | Spanish (Test de Vocabulario en Imágenes Peabody; TVIP); English (Peabody Picture Vocabulary Test; PPVT-5) | Word Reading Accuracy; Text Reading Comprehension | Spanish (Letter Word Identification and Passage Comprehension subtests of the Batería III Woodcock-Muñoz Test of Achievement); English (Letter Word Identification and Passage Comprehension subtests of the Woodcock-Johnson IV Test of Achievement) |  |  |  |
|  |  | Morphological Awareness | Within; Across (Spanish-English) | Spanish (Early Lexical Morphology Measure – Spanish; ELMM-S); English (Early Lexical Morphology Measure; ELMM) | Word Reading Accuracy; Text Reading Comprehension | Spanish (Letter Word Identification and Passage Comprehension subtests of the Batería III Woodcock-Muñoz Test of Achievement); English (Letter Word Identification and Passage Comprehension subtests of the Woodcock-Johnson IV Test of Achievement) |  |  |  |
|  |  | Word Decoding | Within; Across (Spanish-English) | Spanish (Letter Word Identification subtest of the Batería III Woodcock-Muñoz Test of Achievement); English (Letter Word Identification subtest of the Woodcock-Johnson IV Test of Achievement) | Text Reading Comprehension | Spanish (Passage Comprehension subtest of the Batería III Woodcock-Muñoz Test of Achievement); English (Passage Comprehension subtest of the Woodcock-Johnson IV Test of Achievement) |  |  |  |
| **41** | Heterogenous  (English, German, French, Kurdish, Dutch, Turkish, Arabic, or Polish) - Norwegian | Receptive Vocabulary | Within | Norwegian (Vocabulary subtest of the Wechsler Intelligence Scale for Children; WISC-4) | Word Reading Accuracy; Text Reading Comprehension | Norwegian (TOSWRF; NARA) | Grade 5 | 91 | Norway |
|  |  | Oral Language Comprehension | Within | Norwegian (NARA) | Word Reading Accuracy; Text Reading Comprehension | Norwegian (TOSWRF; NARA) |  |  |  |
|  |  | Morphological Awareness | Within | Norwegian (Derivational Morphemes and Non-Words; Knowledge of Conjunctions) | Text Word Reading Accuracy; Text Reading Comprehension | Norwegian (TOSWRF; NARA) |  |  |  |
|  |  | Word Decoding | Within | Norwegian (Word Chain Test; Adaptation of Test of Silent Word Reading Fluency; TOSWRF) | Text Reading Comprehension | Norwegian (NARA) |  |  |  |

*Note.* Studies 1, 2, 8, 12, 16, 23, 28, 34, 35, and 36 assess multiple simultaneous bilingual language groups; ^*^= Studies (7, 15, 19, 22, 23, 24) that only assessed phonemic awareness; Within-Language Testing Medium refers to literacy precursors and outcome measures assessed in the same language; Across-Language Testing Medium refers to literacy precursors and outcome measures assessed in different languages.
