## Supplemental Table 7 for "Early Precursors of Literacy Development in Simultaneous Bilinguals: A Systematic Review and Meta-Analysis"

**S7**

*Precursor-Outcome Associations in Simultaneous Bilingual Children, based on the Type of Literacy Precursor and Reading Outcome Measure Assessed.* Table of significant and non-significant precursor-outcome associations, based on the type of literacy precursor (indicated on the left) and reading outcome measure (indicated on top) assessed – along with the bilingual language combinations spoken by simultaneous bilinguals in each study (as denoted by *). S7 indicates significant and non-significant associations between given literacy precursors (indicated on the left) and given reading outcome measures (indicated on top), as reported by the individual studies listed in S4.

|  | **Word Reading Accuracy** | | **Word Reading Fluency** | | **Non-Word Reading Accuracy** | | **Non-Word Reading Fluency** | | **Text Reading Accuracy** | | **Text Reading Fluency** | | **Text Reading Comprehension** | | | |
| --- | --- | --- | --- | --- | --- | --- | --- | --- | --- | --- | --- | --- | --- | --- | --- | --- |
| **Literacy Precursors**  (n=number of studies) | *Significant* | *Not Significant* | *Significant* | *Not Significant* | *Significant* | *Not Significant* | *Significant* | *Not Significant* | *Significant* | *Not Significant* | *Significant* | *Not Significant* | *Significant* | | *Not Significant* | |
| **Phonological Awareness**  (n=19) | 1*Chinese-English;  2*Chinese (dominant)-English and English (dominant)-Chinese;  3*Chinese-English;  4*Heterogeneous-English;  ^+^7*French-English, and Other-English;  8*Heterogeneous (Malay; Tamil; Mandarin Chinese)-English  9*Russian-Hebrew;  14*Hindi-English;  ^+^15*Spanish-English (Spanish);  ^+^22*Welsh-English  +23*Chinese-English;  32*Chinese-English;  33*Chinese-English | ^+^15*Spanish-English (French) |  | ^+^15*Spanish-English (French & Spanish) | 9*Russian-Hebrew;  14*Hindi-English;  ^+^15*Spanish-English (French & Spanish);    33*Chinese-English |  |  | ^+^15*Spanish-English (French & Spanish) | 9*Russian-Hebrew;  +15*(Spanish-English (French & Spanish) |  | 9*Russian-Hebrew;    ^+^19*Mijikenda; Kikamba-Kiswahili | ^+^15*Spanish-English (French & Spanish) | 23*Chinese-English  ^+^24*Turkish-German;  33*Chinese-English | |  | |
|  | 35*Spanish-English and Chinese-English;  38*Chinese-English  40*Spanish-English |  |  |  |  |  |  |  |  |  |  |  | 40*Spanish-English | | 36*Heterogeneous(dominant)-German and German (dominant)-Heterogenous | |
| **Letter Knowledge**  (n=2) | 4*Heterogeneous-English;  13*Maya-Spanish |  |  |  |  |  |  |  |  |  |  |  |  | |  | |
| **RAN/Serial Recall**  (n=6) | 4*Heterogeneous-English;  22*Welsh-English;  33*Chinese-English;  38*Chinese-English |  |  |  | 33*Chinese-English |  |  |  |  |  |  |  | 33*Chinese-English | | 26*Heterogenous-Dutch;  36*Heterogeneous(dominant)-German and German (dominant)-Heterogenous | |
| **Oral Language/Listening Comprehension**  (n=4) | 10*Spanish-English;  11*Spanish-English;  16*Logographic-English and Alphabetic-English | 41*Heterogenous-Norwegian |  |  | 16*Logographic-English and Alphabetic-English |  |  |  |  |  |  |  | 11*Spanish-English;  16*Logographic-English and Alphabetic-English ;  41*Heterogenous-Norwegian | |  | |
| **Receptive Vocabulary**  (n=21) | 1*Chinese-English;  3*Chinese-English;  8*Heterogeneous (Malay; Tamil; Mandarin Chinese)-English  10*Spanish-English;  11*Spanish-English;  16*Logographic-English and Alphabetic-English;      22*Welsh-English;  +23*Chinese-English  25*Welsh-English;  33*Chinese-English;  35*Spanish-English and Chinese-English;  37*Heterogeneous-English;  38*Chinese-English;  39*Urdu-English;  40*Spanish-English;  41*Heterogenous-Norwegian | 30*Turkish-Dutch |  |  | 16*Logographic-English and Alphabetic-English;            33*Chinese-English |  |  |  | 25*Welsh-English |  | ^+^19*Mijikenda; Kikamba-Kiswahili |  | 11* Spanish-English;  16*Logographic-English and Alphabetic-English;  21*Urdu-Norwegian;  23*Chinese-English;  25*Welsh-English;  26*Heterogenous-Dutch  30*Turkish-Dutch;  36*Heterogeneous(dominant)-German and German (dominant)-Heterogenous;  37*Heterogeneous-English;  39*Urdu-English;  40*Spanish-English;  41*Heterogenous-Norwegian            33*Chinese-English |  | | |
| **Expressive Vocabulary**  (n=5) | 6*Spanish-English;  33*Chinese-English;  38*Chinese-English;  39*Urdu-English |  |  |  | 33*Chinese-English |  |  |  |  |  |  |  | 24*Turkish-German ;  33*Chinese-English;  39*Urdu-English |  | | |
| **Syntactic Awareness**  (n=5) | 4*Heterogeneous-English;  17*Italian-English (Italian and English);  37*Heterogeneous-English | 33*Chinese-English |  |  | 17*Italian-English  (Italian) | 17*Italian-English  (English);  33*Chinese-English |  |  |  |  |  |  | 33*Chinese-English; 36*Heterogeneous(dominant)-German and German (dominant)-Heterogenous;37*Heterogeneous-English |  | | |
| **Morphological Awareness**  (n=11) | 3*Chinese-English;  5*Malay-English  +23*Chinese-English;  33*Chinese-English;  35*Spanish-English and Chinese-English;  37*Heterogeneous-English;  38*Chinese-English;  40*Spanish-English;  41*Heterogenous-Norwegian | 1*Chinese-English |  | 5*Malay-English | 33*Chinese-English |  |  |  |  |  |  |  | +23*Chinese-English;  33*Chinese-English;  36*Heterogeneous(dominant)-German and German(dominant)-Heterogenous;  37*Heterogeneous-English;  40*Spanish-English;  41*Heterogenous-Norwegian |  | | |
| **Working Memory**  (n=5) | 17*Italian-English (Italian) | 17*Italian-English (English);  31*Chinese-English;  33*Chinese-English;  39*Urdu-English |  |  |  | 17*Italian-English (Italian and English);  33*Chinese-English |  |  |  |  |  |  | 33*Chinese-English;  39*Urdu-English | 36*Heterogeneous(dominant)-German and German (dominant)-Heterogenous | | |
| **Verbal Short-Term Memory**  (n=1) |  | 4*Heterogeneous-English |  |  |  |  |  |  |  |  |  |  |  |  | | |
| **Non-Verbal Intelligence**  (n=8) | 22*Welsh-English;  32*Chinese-English;  37*Heterogeneous-English;  38*Chinese-English | 31*Chinese-English;  33*Chinese-English;  39*Urdu-English |  |  |  | 33*Chinese-English |  |  |  |  |  |  | 3*Chinese-English;  37*Heterogeneous-English | 39*Urdu-English | | |
| **Word/Non-Word Decoding**  (n=10) |  |  |  |  |  |  |  |  |  |  |  |  | 21*Urdu-Norwegian;  23*Chinese-English;  24*Turkish-German  30*Turkish-Dutch;  33*Chinese-English;  37*Heterogeneous-English;  39*Urdu-English;  40*Spanish-English;  41*Heterogenous-Norwegian | | | 26*Heterogenous-Dutch |
| **Semantic Awareness**  (n=3) |  | 7*French-English;  9*Russian-Hebrew |  |  |  | 9*Russian-Hebrew |  |  |  | 9*Russian-Hebrew |  | 9*Russian-Hebrew |  | | | 26*Heterogenous-Dutch |
| **Spelling**  (n=3) | 4*Heterogeneous-English;  17*Italian-English (Italian and English);  38*Chinese-English |  |  |  | 17*Italian-English (Italian and English) |  |  |  |  |  |  |  |  | | |  |
| **VA Span**  (n=2) | 15*Spanish-English (French & Spanish);    34* French-Basque | 34*Spanish-Basque | 15*Spanish-English (French & Spanish);  34* French-Basque | 34*Spanish-Basque | 15*Spanish-English (French & Spanish);    34*French-Basque | 34*Spanish-Basque | 15*Spanish-English (French & Spanish);    34* French-Basque | 34*Spanish-Basque | 15*Spanish-English (French) | 15* Spanish- English (Spanish) | 15* Spanish-English (French & Spanish) |  |  | | |  |
| **Orthographic Processing**  (n=2) | 2* Younger English (dominant)-Chinese and Older Chinese (dominant)-English;  38*Chinese-English | 2*Younger Chinese (dominant)-English and Older English (dominant)-Chinese |  |  |  |  |  |  |  |  |  |  |  | | |  |
| **Environmental Print Awareness**  (n=1) | 4*Heterogeneous-English |  |  |  |  |  |  |  |  |  |  |  |  | | |  |
| **Name Writing**  (n=1) | 13*Maya-Spanish |  |  |  |  |  |  |  |  |  |  |  |  | | |  |
| **Sub-Lexical/Phonological Speech Perception Task** (n=1) |  | 28*Basque-Spanish | 28*Basque-Spanish |  |  | 28*Basque-Spanish | 28*Basque-Spanish |  |  | 28*Basque-Spanish | 28*Basque-Spanish |  |  | | |  |
| **Sentence Priming Task** (n=1) |  |  |  |  |  |  |  |  | 29*Hebrew-English |  | 29*Hebrew-English |  |  | | | 29*Hebrew-English |
| **Novel Word Learning**  (n=1) |  | 39*Urdu-English |  |  |  |  |  |  |  |  |  |  | 39*Urdu-English | | |  |

*Note.** = specifies type of bilingual language background; ^+^= Studies (7, 15, 19, 22, 23, 24) that only assessed phonemic awareness.
