## Supplemental Table 8 for "Early Precursors of Literacy Development in Simultaneous Bilinguals: A Systematic Review and Meta-Analysis"

**S8**

*Precursor-Outcome Associations Assessed in One Language for Simultaneous Bilingual Children.* Table of significant and non-significant within- and cross-language precursor-outcome associations for the literacy precursors (indicated on the left, along with number of studies assessed) and/or reading outcome measures assessed in one bilingual language – along with the language(s) of assessment (as denoted by ^PL^ and ^OL^). S8 indicates significant and non-significant associations between given literacy precursors (indicated on the left) and reading outcome measures, as reported by the individual studies listed in S4.

| **Literacy Precursors** (n=number of studies) | ***Literacy Precursors and Outcomes Assessed in One Language*** | | | ***Literacy Precursors Assessed in Both Languages and Outcomes Assessed in One Language*** | | ***Literacy Precursors Assessed in One Language and* Outcomes *Assessed in Both Languages*** | |
| --- | --- | --- | --- | --- | --- | --- | --- |
|  | *Significant*  *Within-Language* | *Significant*  *Cross-Language* | *Not Significant*  *(Within-Language)* | *Significant Within-Language* | *Significant Cross-Language* | *Significant Within-Language* | *Significant Cross-Language* |
| **Phonological Awareness** (n=13) | 2 **^PL & OL^** (English);  4 **^PL & OL^** (English); ^+^7(Heterogenous-English bilinguals)  **^PL & OL^** (English);  8 **^PL & OL^** (English);  9 **^PL & OL^** (Hebrew);  ^+^19 **^PL & OL^** (Kiswahili);  +23 **^PL & OL^** (English)  ^+^24 **^PL & OL^** (German) |  | 36 Heterogeneous(dominant)-German and German (dominant)-Heterogenous  **^PL & OL^** (German) | 1 **^PL^** (Chinese and English)  **^OL^** (English);  32 **^PL & OL^** (Chinese and English) | 1 **^PL^** (Chinese and English)  **^OL^** (English);  32 **^PL^** (Chinese and English)  **^OL^** (English);  35 **^PL^** (Spanish and English; Chinese and English)  **^OL^** (English) |  | 38 **^PL & OL^**(English and Chinese) |
| **Letter Knowledge**  (n=1) | 4 **^PL & OL^** (English) |  |  |  |  |  |  |
| **RAN/Serial Recall** (n=4) | 4 **^PL & OL^** (English) |  | 26 **^PL & OL^** (Dutch);  36 Heterogeneous(dominant)-German and German (dominant)-Heterogenous  **^PL & OL^** (German) |  |  | 22 **^PL & OL^** (Welsh and English) | 22 **^PL & OL^** (Welsh and English) |
| **Oral Language/Listening Comprehension** (n=2) | 16 **^PL & OL^** (English);  41 **^PL & OL^** (Norwegian) |  |  |  |  |  |  |
| **Receptive Vocabulary** (n=14) | 1 **^PL & OL^** (English);  16 **^PL & OL^** (English);  19 **^PL & OL^** (Kiswahili);  23 **^PL & OL^** (English);  26 **^PL & OL^** (Dutch);  36 Heterogeneous(dominant)-German and German (dominant)-Heterogenous  **^PL & OL^** (German);  37 **^PL & OL^** (English);  39 **^PL & OL^** (English);  41 **^PL & OL^** (Norwegian) |  |  | 8 **^PL^** (Malay, Tamil or Mandarin-Chinese and English)  **^OL^** (English);  21 **^PL^** (Norwegian and Urdu)  **^OL^** (Norwegian)  30 **^PL^** (Turkish and Dutch)  **^OL^** (Dutch) | 8 **^PL^** (Malay, Tamil or Mandarin-Chinese and English)  **^OL^** (English);  21 **^PL^** (Norwegian and Urdu)  **^OL^** (Norwegian);  35 **^PL^** (Spanish and English; Chinese and English)  **^OL^** (English) | 22 **^PL^** (Welsh)  **^OL^** (Welsh and English) | 22 **^PL^** (Welsh)  **^OL^** (Welsh and English) |
| **Expressive Vocabulary** (n=3) | 24 **^PL & OL^** (German);  39 **^PL & OL^** (English) |  |  |  | 6 **^PL^** (Spanish and English)  **^OL^** (English) |  |  |
| **Syntactic Awareness** (n=3) | 4 **^PL & OL^** (English);  36 Heterogeneous(dominant)-German and German (dominant)-Heterogenous  **^PL & OL^** (German);  37 **^PL & OL^** (English) |  |  |  |  |  |  |
| **Morphological Awareness** (n=7) | 23 **^PL & OL^** (English);  36 Heterogeneous(dominant)-German and German (dominant)-Heterogenous  **^PL & OL^** (German);  37 **^PL & OL^** (English);  41 **^PL & OL^** (Norwegian) |  | 1 **^PL^** (Chinese)  **^OL^** (English)  ^^^ns across language associations (different languages assessed for literacy precursor and outcome measure) |  | 35 **^PL^** (Spanish and English; Chinese and English)  **^OL^** (English) | 5 **^PL^** (English)  **^OL^** (Malay and English) | 5 **^PL^** (English)  **^OL^** (Malay and English) |
| **Working Memory** (n=3) | 39 **^PL & OL^** (English) |  | 31 **^PL & OL^** (English);  36 Heterogeneous(dominant)-German and German (dominant)-Heterogenous  **^PL & OL^** (German) |  |  |  |  |
| **Verbal Short-Term Memory** (n=1) |  |  | 4 **^PL & OL^** (English) |  |  |  |  |
| **Non-Verbal Intelligence** (n=5) | 32 **^OL^** (English);  37 **^PL & OL^** (English) | 39 **^PL & OL^** (English) | 31 **^OL^** (English) |  |  | 22 **^OL (^**Welsh and English); | 22 **^OL^** (Welsh and English) |
| **Word/Non-Word Decoding** (n=8) | 21 **^PL & OL^ (**Norwegian);  23 **^PL & OL^** (English);  24 **^PL & OL^** (German);  30 **^PL & OL^** (Dutch);  37 **^PL & OL^** (English);  39 **^PL & OL^** (English);  41 **^PL & OL^** (Norwegian) |  | 26 **^PL & OL^** (Dutch) |  |  |  |  |
| **Semantic Awareness** (n=3) |  |  | 7(Heterogenous-English bilinguals)  **^PL & OL^** (English);  9 **^PL & OL^** (Hebrew);  26 **^PL & OL^** (Dutch) |  |  |  |  |
| **Spelling** (n=1) | 4 **^PL & OL^** (English) |  |  |  |  |  |  |
| **VA Span** (n=1) |  |  |  |  | 34 **^PL^** (Spanish, French, Basque)  **^OL^** (Basque) |  |  |
| **Orthographic Processing** (n=1) | 2 **^PL & OL^** (English); |  |  |  |  |  |  |
| **Environmental Print Awareness** (n=1) | 4 **^PL & OL^** (English) |  |  |  |  |  |  |
| **Sub-Lexical/Phonological Speech Perception Task** (n=1) | 28 **^PL & OL^** (Spanish) |  |  |  |  |  |  |
| **Sentence Priming Task** (n=1) |  | 29 **^PL & OL^**(Hebrew)  ^^^Different languages assessed for literacy precursor and outcome measure |  |  |  |  |  |
| **Novel Word Learning** (n=1) | 39 **^PL & OL^** (English) |  |  |  |  |  |  |

*Note.* ^^^ = specifies additional information; ns= not significant; ^+^= Studies (7, 19, 23, 24) that only assessed phonemic awareness; ^PL^= (precursor language) indicates language assessed for literacy precursor measures; ^OL^= (outcome language) indicates language assessed for reading outcome measures. The term *within-language precursor-outcome associations* refer to associations between literacy precursors and reading outcomes assessed in the same language. The term *cross-language precursor-outcome* associations refer to precursors and outcomes assessed in two different languages.
