## Supplemental Table 9 for "Early Precursors of Literacy Development in Simultaneous Bilinguals: A Systematic Review and Meta-Analysis"

**S9**

*Precursor-Outcome Associations Assessed in Both Bilingual Languages for Simultaneous Bilingual Children.* Table of significant and non-significant within- and cross-language precursor-outcome associations for the literacy precursors (indicated on the left, along with number of studies assessed) and reading outcome measures assessed in both bilingual languages – along with the language(s) of assessment (as denoted by ^PL^ and ^OL^). S9 indicates significant and non-significant associations between given literacy precursors (indicated on the left) and reading outcomes, as reported by the individual studies listed in S4.

| **Literacy Precursors** (n=number of studies) | ***Literacy Precursors and Outcomes Assessed in Both Bilingual Languages*** | | | | |  |
| --- | --- | --- | --- | --- | --- | --- |
|  | ***Significant Within-Language for One Language*** | ***Significant Within-Language for Both Languages*** | ***Significant Cross-Language for One Language*** | ***Significant Cross-Language for Both Languages*** | ***Not* *Significant***  ***(Cross-Language)*** | ***Not* *Significant***  ***(Both Within- and Cross-Language)*** |
| **Phonological Awareness** (n=9) | ^+^7(French-English bilinguals)  **^PL & OL^** (French and ^English);  33 **^PL & OL^**(*English and Chinese);  35 **^PL & OL^**(*English);  38 **^PL & OL^**(English and *Chinese) | 3 **^PL & OL^** (Chinese and English);  14 **^PL^** (neural study) **^& OL^** (Hindi and English);  ^+^15 **^PL & OL^** (Spanish and French);  ^+^22 **^PL & OL^** (Welsh and English);  40 **^PL & OL^**(Spanish and English) |  | ^+^15 **^PL & OL^** (Spanish and French);  ^+^22 **^PL & OL^** (Welsh and English);  40 **^PL & OL^**(Spanish and English) | 3 **^PL & OL^** (Chinese and English);  33 **^PL & OL^** (English and Chinese) |  |
| **Letter Knowledge**  (n=1) |  | 13 **^PL & OL^** (Maya and Spanish) |  | 13 **^PL & OL^** (Maya and Spanish) |  |  |
| **RAN/Serial Recall**  (n=2) |  | 38 **^PL & OL^**(English and Chinese) | 33 **^PL & OL^**(English and Chinese)  *Chinese serial recall – English reading |  |  |  |
| **Oral Language/Listening Comprehension** (n=2) |  | 10 **^PL & OL^** (Spanish and English);  11 **^PL & OL^** (Spanish and English) |  | 10 **^PL & OL^** (Spanish and English);  11 **^PL & OL^** (Spanish and English) |  |  |
| **Receptive Vocabulary** (n=8) | 35 **^PL & OL^**(*English) | 3 **^PL & OL^** (Chinese and English);  10 **^PL & OL^** (Spanish and English);  11 **^PL & OL^** (Spanish and English);  25 **^PL & OL^** (Welsh and English);  33 **^PL & OL^**(English and Chinese);  38 **^PL & OL^**(English and Chinese);  40 **^PL & OL^**(Spanish and English) |  | 10 **^PL & OL^** (Spanish and English);  11 **^PL & OL^** (Spanish and English);  40 **^PL & OL^**(Spanish and English) | 3 **^PL & OL^** (Chinese and English);  33 **^PL & OL^**(English and Chinese) |  |
| **Expressive Vocabulary** (n=2) |  | 33 **^PL & OL^**(English and Chinese);  38 **^PL & OL^**(English and Chinese) | 33 **^PL & OL^**(English and Chinese)  *English vocabulary – Chinese reading |  |  |  |
| **Syntactic Awareness** (n=2) | 33 **^PL & OL^**(*English and Chinese) | 17 **^PL & OL^** (Italian and English) | 17 **^PL & OL^** (Italian and English)  *English syntactic awareness- Italian reading |  | 33 **^PL & OL^**(English and Chinese) |  |
| **Morphological Awareness** (n=5) | 35 **^PL & OL^**(*English) | 3 **^PL & OL^** (Chinese and English);  33 **^PL & OL^**(English and Chinese);  38 **^PL & OL^**(English and Chinese);  40 **^PL & OL^**(Spanish and English) |  | 40 **^PL & OL^**(Spanish and English) | 3 **^PL & OL^** (Chinese and English);  33 **^PL & OL^**(English and Chinese) |  |
| **Working Memory**  (n=2) |  | 33 **^PL & OL^**(English and Chinese) | 17 **^PL & OL^** (Italian and English)  *English working memory- Italian reading;  33 **^PL & OL^**(English and Chinese)  * Chinese working memory- English reading |  |  |  |
| **Non-Verbal Intelligence** (n=2) | 33 **^PL & OL^**(*English and Chinese) | 38*Chinese-English | 33 **^PL & OL^**(English and Chinese)  *Chinese non-verbal intelligence vocabulary – English reading |  |  |  |
| **Word/Non-Word Decoding** (n=2) |  | 33 **^PL & OL^**(English and Chinese);  40 **^PL & OL^**(Spanish and English) |  | 40 **^PL & OL^**(Spanish and English) | 33 **^PL & OL^**(English and Chinese) |  |
| **Semantic Awareness** (n=1) |  |  |  |  |  | 7(French-English bilinguals)  **^PL & OL^** (French and English) |
| **Spelling** (n=2) |  | 17 **^PL & OL^** (Italian and English);  38*Chinese-English |  | 17 **^PL & OL^** (Italian and English) |  |  |
| **VA Span** (n=1) |  | 15 **^PL & OL^** (Spanish and French) |  | 15 **^PL & OL^** (Spanish and French) |  |  |
| **Orthographic Processing** (n=1) |  | 38 **^PL & OL^**(English and Chinese) |  |  |  |  |
| **Name Writing** (n=1) | 13 **^OL^** (*Maya and Spanish) |  |  |  |  |  |

*Note.*^+^= Studies (7, 15, 22) that only assessed phonemic awareness; ^PL^= (precursor language) indicates language assessed for literacy precursor measures; ^OL^= (outcome language) indicates language assessed for reading outcome measures; ^= literacy precursor assessed in both bilingual languages, but only statistically analyzed in one bilingual language; *= language(s) with significant precursor-outcome associations, for precursors assessed in both languages but only significant in one language. The term *significant within-language precursor-outcome associations for one language* refers to literacy precursor and reading outcome measures assessed in the same language and significant in one language. The term *significant within-language precursor-outcome associations for both languages* refers to measures assessed in the same language and significant in both languages. The term *significant across-language precursor-outcome associations for literacy outcomes in one language* refers to precursor and outcome measures assessed in different languages, and significant for outcomes in one language. The term *significant cross-language precursor-outcome associations for literacy outcomes in both languages* refers to precursor and outcome measures assessed in different languages, and significant for outcomes in both languages.
