## Supplemental Figure 10 for "Early Precursors of Literacy Development in Simultaneous Bilinguals: A Systematic Review and Meta-Analysis"

**S10**

*Meta-Analysis for Phonological Awareness in relation to Word and Non-Word Reading (English language only)*. Forestplot indicating correlational effect size for random-effects model (r= 0.5068; 95% CI: 0.4055, 0.5958; p< 0.0001) and individual correlations/study between phonological awareness and word/non-word reading, for within-language studies that only assessed phonological awareness and word/non-word reading measures in English.

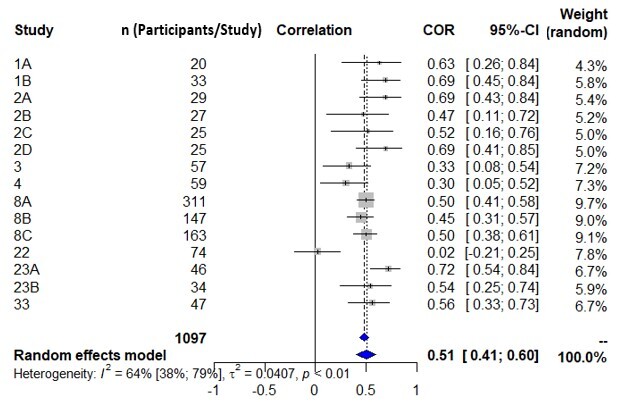
