## Supplemental Figure 11 for "Early Precursors of Literacy Development in Simultaneous Bilinguals: A Systematic Review and Meta-Analysis"

**S11**

*Meta-Analysis for Phonological Awareness in relation to Word and Non-Word Reading (Another heritage or societal language only)*. Forestplot indicating correlational effect size for random-effects model (r= 0.3563; 95% CI: 0.1169, 0.5569; p=0.0042) and individual correlations/study between phonological awareness and word/non-word reading, for within-language studies that only assessed phonological awareness and word/non-word reading measures in another (non-English) heritage or societal language.

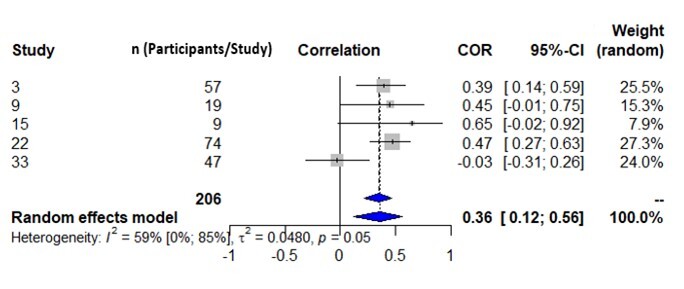
