## Supplemental Figure 12 for "Early Precursors of Literacy Development in Simultaneous Bilinguals: A Systematic Review and Meta-Analysis"

**S12**

*Meta-Analysis for Vocabulary in relation to Word and Non-Word Reading (English language only)*. Forestplot indicating correlational effect size for random-effects model (r= 0.4027; 95% CI: 0.2862, 0.5075; p< 0.0001) and individual correlations/study between vocabulary and word/non-word reading, for within-language studies that only assessed vocabulary and word/non-word reading measures in English.

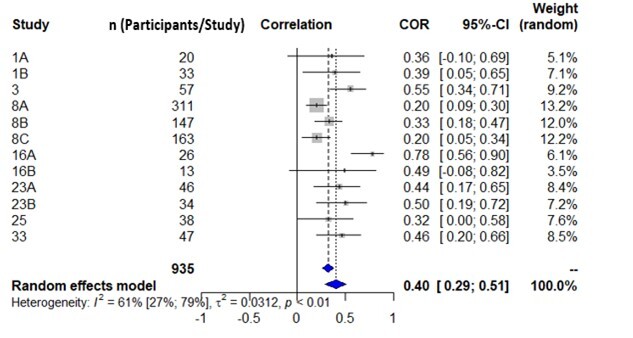
