## Supplemental Figure 13 for "Early Precursors of Literacy Development in Simultaneous Bilinguals: A Systematic Review and Meta-Analysis"

**S13**

*Meta-Analysis for Vocabulary in relation to Word and Non-Word Reading (Another heritage or societal language only)*. Forestplot indicating correlational effect size for random-effects model (r= 0.3133; 95% CI: 0.1383, 0. 4692; p< 0.0006) and individual correlations/study between vocabulary and word/non-word reading, for within-language studies that only assessed vocabulary and word/non-word reading measures in another (non-English) heritage or societal language.

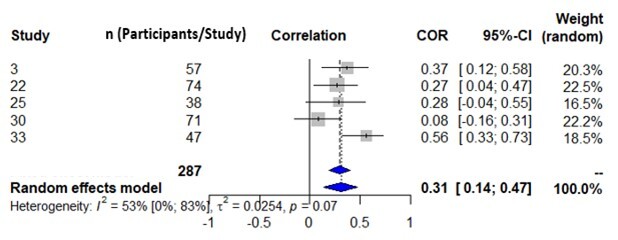
