## Supplemental Figure 14 for "Early Precursors of Literacy Development in Simultaneous Bilinguals: A Systematic Review and Meta-Analysis"

**S14**

*Meta-Analysis for Morphological Awareness in relation to Word and Non-Word Reading*. Forestplot indicating correlational effect size for random-effects model (r= 0.5005; 95% CI: 0.1441, 0.7420; p=0.0078) and individual correlations/study between morphological awareness and word/non-word reading.

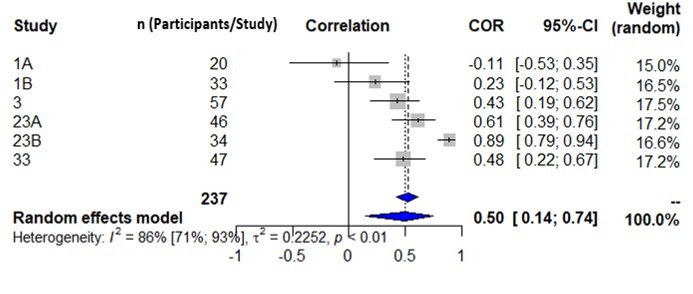
