## Supplemental Figure 15 for "Early Precursors of Literacy Development in Simultaneous Bilinguals: A Systematic Review and Meta-Analysis"

**S15**

*Meta-Analysis for Vocabulary in relation to Text Reading Comprehension*. Forestplot indicating correlational effect size for random-effects model (r= 0.5706; 95% CI: 0.3669, 0.7221; p< 0.0001) and individual correlations/study between vocabulary and text reading comprehension.

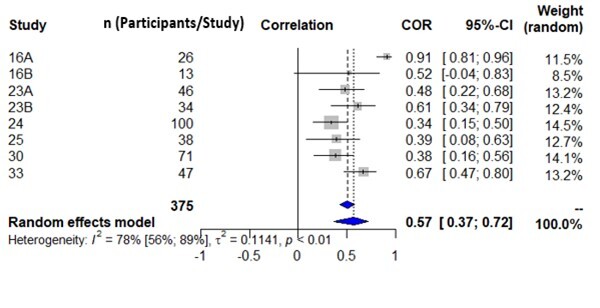
