## Supplemental Figure 16 for "Early Precursors of Literacy Development in Simultaneous Bilinguals: A Systematic Review and Meta-Analysis"

**S16**

*Meta-Analysis for Word/Non-Word Decoding in relation to Text Reading Comprehension*. Forestplot indicating correlational effect size for random-effects model (r= 0.6741; 95% CI: 0.3705, 0.8476; p=0.0002) and individual correlations/study between word/non-word decoding and text reading comprehension.

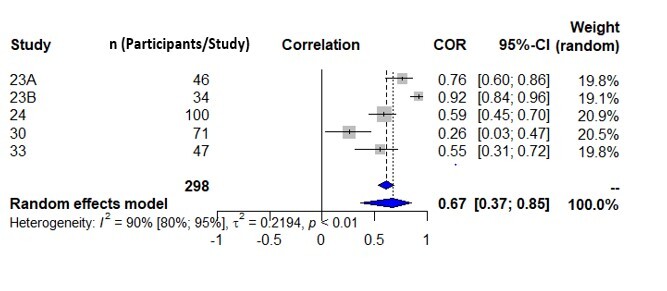
