## Supplemental Figure 17 for "Early Precursors of Literacy Development in Simultaneous Bilinguals: A Systematic Review and Meta-Analysis"

**S17**

*Subgroup Analysis for Phonological Awareness in relation to Word/Non-Word Reading, based on Type of Literacy Precursor and Outcome Measure Testing.* Forestplot indicating correlational effect size for random-effects model (test for overall effect: r= 0.4713; 95% CI: 0.3850, 0.5494; p<0.0001) and individual correlations/study between phonological awareness and word/non-word reading, based on type of testing subgroups (i.e., whether phonological awareness and word/non-word reading measures were assessed in the same language [within-language testing subgroup; r= 0.5031; 95% CI: 0.4098, 0.5860] or different language [cross-language testing subgroup; r= 0.3485; 95% CI: 0.1577, 0.5142]; test for subgroup differences: Q=2.48; p=0.1154).

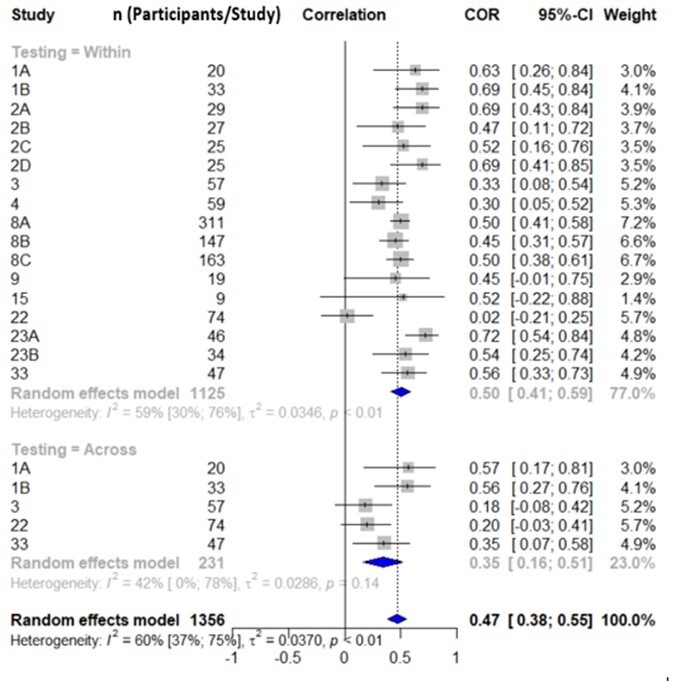
