## Supplemental Figure 18 for "Early Precursors of Literacy Development in Simultaneous Bilinguals: A Systematic Review and Meta-Analysis"

**S18**

*Subgroup Analysis for Vocabulary in relation to Word/Non-Word Reading, based on Type of Literacy Precursor and Outcome Measure Testing.* Forestplot indicating correlational effect size for random-effects model (test for overall effect: r= 0.3254; 95% CI: 0.2135, 0.4289; p<0.0001) and individual correlations/study between vocabulary and word/non-word reading, based on type of testing subgroups (i.e., whether vocabulary and word/non-word reading measures were assessed in the same language [within-language testing subgroup; r= 0.4221; 95% CI: 0.28889, 0.5392] or different language [cross-language testing subgroup; r= 0.2026; 95% CI: 0.0644, 0.3332]; test for subgroup differences: Q=5.32; p=0.0210).

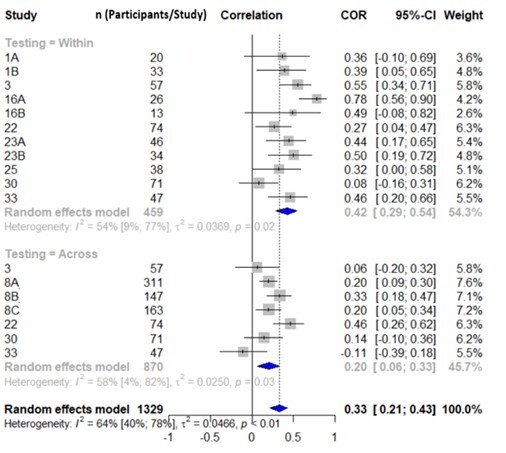
